# Within-host pathogen population diversity predicts treatment response in Tuberculosis

**DOI:** 10.64898/2026.06.16.26355818

**Authors:** Sanjana G. Kulkarni, Maximillian G. Marin, Brendon C. Mann, Noorjahn Rawoot, Suchitra Goodwin, Nina Cesare, Robin M. Warren, Karen R. Jacobson, Maha R. Farhat

## Abstract

**Background:** Tuberculosis (TB) treatment outcomes remain suboptimal, and standard clinical diagnostics cannot reliably identify patients at high risk of treatment failure or relapse at the time of diagnosis. While within-host *Mycobacterium tuberculosis* genetic diversity is hypothesized to reflect the viable bacterial burden and adaptive capacity of the infection, its clinical prognostic value remains unknown.

**Methods:** We conducted a prospective cohort study of 364 patients with newly diagnosed, rifampicin-susceptible pulmonary TB in South Africa. Patients received standard 6-month therapy and were monitored for up to two years to ascertain composite unfavorable outcomes (treatment failure, death, or relapse). To accurately detect low-frequency (unfixed) genetic variants and eliminate reference bias artifacts, we mapped medium to high depth short-read sequences against matched, patient-specific long-read assemblies. The association between baseline pathogen genetic diversity and clinical outcomes was evaluated using multivariable Cox proportional-hazards models.

**Results:** After bioinformatic filtering, true unfixed variants were relatively rare but significantly enriched in genes mediating pathogen adaptation and drug tolerance, including transporter proteins and two-component regulatory systems. Within-host bacterial genetic diversity (*i.e.,* the total number of unfixed variants) ranged from 0–20, with a median of 1 per patient. In survival analysis adjusting for known clinical risk factors—including HIV status, prior TB, baseline smear positivity, and radiographic lung involvement—baseline within-host genetic diversity emerged as a strong, independent predictor of unfavorable treatment outcomes. For patients with greater than 3 unfixed variants at diagnosis, each increase of 5 unfixed variants was associated with more than double the risk of a composite unfavorable outcome (adjusted Hazard Ratio, 2.36; 95% CI, 1.27 to 4.39; p=0.007).

**Conclusions:** Baseline within-host pathogen genetic diversity is an independent predictor of unfavorable TB treatment outcomes. As sequencing becomes increasingly integrated into routine diagnostics, quantifying unfixed variants is an accessible approach that promises to risk-stratify patients and guide the duration of individualized regimens.

## Introduction

Tuberculosis (TB) treatment outcomes remain suboptimal. In routine programmatic settings, up to 15% of patients with drug-susceptible TB experience treatment failure or death,^1,2^ with post-treatment relapse affecting a comparable proportion. Outcomes for drug-resistant TB are even worse, with global success rates stalling near 60% and combined failure and mortality approaching 50% in highly resistant cohorts.^2–4^ Crucially, poor outcomes persist even under the strict observation of clinical trials, where 9-14% of patients relapse after standard 6-month therapy or shortened 4-month fluoroquinolone regimens.^5^ These outcomes highlight a critical clinical gap: standard microbiological assessment at treatment initiation cannot identify poor responders. Microscopic metrics of bacterial burden such as smear grade are predictive of outcome but are not accurate and are not currently used clinically to guide treatment composition or duration.^4,6^ These metrics have been criticized for their inherent inability to distinguish live from dead *M. tuberculosis* bacteria. Other metrics of bacterial viability such as transcriptomics are promising but only as follow up metrics measuring response after days to weeks of treatment.^7^ To improve outcomes and accelerate clinical trials of shortened regimens, there is a critical need for a novel microbiological biomarker measurable at treatment initiation that can accurately predict severity of bacterial infection. Because bacterial sequencing is now routinely used for drug resistance diagnosis in many TB care settings and was recently endorsed by the WHO for this indication,^8^ pathogen genetic data will be readily available on a large number of patients in routine clinical care. In this work we hypothesized that pathogen sequencing data can be used for assessing the severity of bacterial infection and predicting treatment response.

Although active tuberculosis typically originates from the transmission of a small infectious inoculum, considerable within-host diversification occurs as the disease progresses. Studies utilizing barcoded *M. tuberculosis* strains in non-human primate models have demonstrated that this diversification is driven largely by genetic drift and spatial compartmentalization. As bacteria seed independent, discontinuous anatomical niches across the lung parenchyma and regional lymphatics, distinct subpopulations evolve independently but can be mixed when cavitary lung disease breaks down.^9^ This evolutionary process is compounded by the chronicity of TB; clinical presentation is often delayed, with patients frequently harboring active, replicating disease for weeks and usually months prior to diagnosis. Under neutral evolutionary theory, the accumulation of this within-host genetic diversity is directly proportional to the effective population size of the bacteria. Consequently, the degree of pathogen diversification can serve as a high-resolution molecular proxy for the total viable bacterial burden and the overall physiological health of the infecting population—capturing critical biological information that may be missed by microscopic assessments.^10–12^

Beyond neutral drift, within-host diversification generates a reservoir of “unfixed” or low frequency genetic variants that facilitate bacterial adaptation to the host environment. This dynamic microevolution enables the pathogen population to explore the fitness landscape, leading to the emergence of subpopulations with resistance mutations (heteroresistance), or mutations that result in metabolic or other types of drug tolerance.^12–16^ Because these adaptive traits directly influence how well the bacterial population can survive the sudden bottleneck of multi-drug therapy, they may be predictive of clinical severity and treatment response. For example, unfixed variants in genes known to encode fluoroquinolones, bedaquiline, and clofazimine resistance at a frequency of 25-75% substantially increase the sensitivity of resistance diagnosis to these drugs from *M. tuberculosis* (*Mtb*) sequencing compared with variants at higher frequency (>75%).^2,17,18^ Therefore, detecting the emergence of unfixed variants—and their dynamics over time during treatment—holds promise for predicting which infections may be harder to treat under standard therapeutic regimens.^11,12,19^

While small retrospective studies have suggested that high within-strain *Mtb* diversity are associated with worse treatment outcomes,^20^ no prospective studies have evaluated within-host genome diversity as a biomarker for treatment response. Also in prior work, the accurate assessment of within-host diversity has been impeded by the difficulty of reliable variant calling; low-frequency variants are highly susceptible to false positives caused by reference bias and uncertain read mapping in standard short-read sequencing. To overcome these technological barriers, here we leveraged long-read sequencing to assemble highly accurate, patient-specific “personal” pathogen genomes.

We designed and implemented a prospective cohort study of 458 individuals with newly diagnosed, rifampicin-susceptible pulmonary TB in Worcester, South Africa^21^ to study within-host pathogen diversity as a metric of severity of bacterial infection. Participants were treated with a standard 6-month regimen and followed with weekly sputum cultures for the first 12 weeks and at treatment completion and underwent further close monitoring for one year to ascertain relapse. Using non-PCR-based Illumina sequencing (at ≥250x depth) mapped against 172 matched long-read personal genomes, we rigorously quantified low-frequency variation during the first weeks of therapy. We find that the number of unfixed within-host variants at baseline predicts poor treatment outcomes independently of microscopic metrics of burden and known host factors, suggesting its potential as a surrogate endpoint in clinical trials and a tool to guide individualized treatment.

## Results

### Patient population

We recruited 458 participants with a new diagnosis of rifampicin-susceptible TB in Worcester, South Africa from 2017 to 2024.^21^ Participants were majority male (61%), aged on average 37 years, and 27% were co-infected with HIV (**Table 1).**

**Table 1.** Patient baseline characteristics. The second column contains information for all participants with a baseline sequencing sample taken within the first 2 weeks of treatment. The third column contains information for the 284 participants with concordant longitudinal sequencing. 5.6% of individuals had an unfavorable outcome, and the relapse rate is 2.6%, which is consistent with previous empirical estimates of the relapse rate for drug-susceptible TB.^25^

| Variable | Baseline Samples (N = 418) | Longitudinal Samples (N = 284) |
| --- | --- | --- |
| **Male sex** | 253 (61%) | 176 (62%) |
| **Age Range** | 15–77 | 15–77 |
| 15–24 | 87 (21%) | 54 (19%) |
| 25–34 | 94 (22%) | 60 (21%) |
| 35–44 | 109 (26%) | 78 (27%) |
| 45–54 | 97 (23%) | 65 (23%) |
| 55–64 | 24 (5.7%) | 22 (7.7%) |
| 65–77 | 7 (1.7%) | 5 (1.8%) |
| **HIV/CD4 Status** |  |  |
| HIV– | 301 (72%) | 216 (76%) |
| HIV+, CD4 ≥ 200 cells/mm<sup>3</sup> | 69 (17%) | 51 (18%) |
| HIV+, CD4 < 200 cells/mm<sup>3</sup> | 43 (10%) | 16 (5.6%) |
| Missing | 5 (1.1%) | 1 (0.4%) |
| **Diabetes** | 22 (5.3%) | 17 (6.0%) |
| **Cigarette smoking**<br>Yes<br>No<br>Missing | 278 (66.5%)<br>139 (33.3%)<br>1 (0.2%) | 192 (68%)<br>92 (32%)<br>0 (0%) |
| **Body Mass Index (BMI)**<br>< 18<br>18–25<br>> 25 | 230 (55%)<br>168 (40%)<br>20 (4.8%) | 162 (57%)<br>109 (38%)<br>13 (4.6%) |
| **Previous TB disease** | 165 (39%) | 121 (43%) |
| **Smear Positivity**<br>Positive<br>Negative<br>Missing | 304 (73%)<br>112 (27%)<br>2 (0.5%) | 231 (81%)<br>53 (19%)<br>0 (0%) |
| **Smear Grade**<br>No AFB<br>Scanty<br>+<br>++<br>+++<br>Missing | 112 (27%)<br>55 (13%)<br>107 (26%)<br>62 (15%)<br>80 (19%)<br>2 (0.5%) | 53 (19%)<br>36 (13%)<br>88 (31%)<br>47 (16%)<br>60 (21%)<br>0 (0%) |
| **Percent lung involved**<br>Median<br>Interquartile Range<br>Missing | 26%<br>26%<br>14 (3.3%) | 29%<br>25%<br>8 (2.8%) |
| **Lineage Mixing**<br>Clonal ($F2 \leq 0.03$)<br>Mixed ($F2 > 0.03$) | 395 (94.5%)<br>23 (5.5%) | 284 (100%)<br>0 (0%) |
| **Lineage (among clonal)**<br>1<br>2<br>3<br>4 | 1 (0.2%)<br>206 (49%)<br>28 (6.7%)<br>178 (43%) | 0 (0%)<br>148 (52%)<br>15 (5%)<br>121 (42%) |
| **Isoniazid resistance**<br>Resistant<br>Susceptible<br>Missing | 36 (8.6%)<br>364 (87%)<br>18 (4.3%) | 20 (7.0%)<br>257 (90.5%)<br>7 (2.5%) |
| **Sequencing Depth**<br>Median<br>Interquartile Range | 311.5<br>67 | 310<br>70.5 |

Of the 458 participants, 418 had a high-quality baseline culture sample sequenced with short reads within the first 2 weeks of treatment initiation, and 310 had a second sequenced sample at a median of 6 weeks on treatment (**Fig 1a**). Of the 418 participants with baseline sequences, 395 were infected with a single *M. tuberculosis* lineage, and 285 of the 310 patients with longitudinal sampling were infected with a single concordant lineage at both time points (*i.e.,* clonal infections). After excluding one outlier pair (**Methods**), we estimate a mutation fixation rate of 0.66 single nucleotide polymorphisms (SNPs)/genome/year (95% confidence interval = 0.36-1.2) between our longitudinal pairs, which is consistent with prior empirical estimates of the *M. tuberculosis* mutation rate.^22–24^

**Figure 1.**
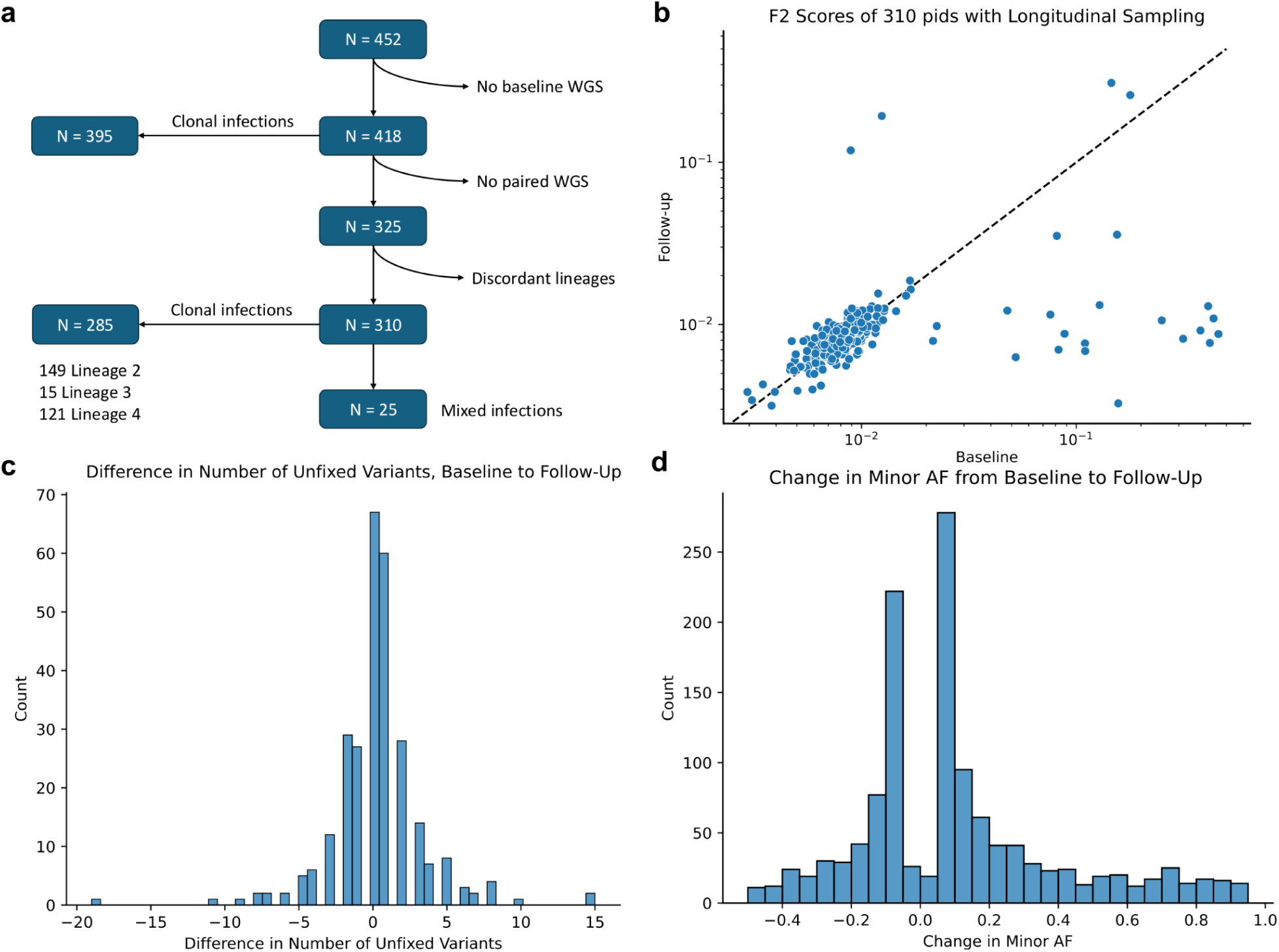
Unfixed genetic variants frequencies in baseline and follow up clinical isolates. **a**: Flow chart of cohort participants and those with single and paired short-read sequencing. **b:** F2 score at baseline (x-axis) and follow-up (y-axis) for 310 participants with at least one overlapping lineage between timepoints. **c:** Distribution of the difference in unfixed variant burden between follow-up and baseline for 284 participants. **d:** Distribution of the difference in minor allele fraction between follow-up and baseline for 1,269 variants.

The remaining 25 patients with sequencing data had a mixed infection in which two or more *M. tuberculosis* lineages were simultaneously isolated at one or both time points. The majority of the participants with mixed lineage infection lost one of the lineages and had persistence of the second lineage in interval follow up (**Supplementary Fig. 4, Supplementary Table 1**).

### Unfixed genetic variant calls are error prone and are relatively rare in culture isolates

Given that calling unfixed variants is highly prone to reference bias,^26^ we trained a logistic regression model to predict the probability of error for a given unfixed variant using hybrid short-and long-read personal genomes from 172 isolates (**Methods**, **Supplementary Results**). On the training data (N = 802 variant calls: 401 real, 401 not real), the model achieved an area under the curve (AUC) of 0.991, precision of 0.978, and recall of 0.978 for calling variants with allele frequency (AF) between 5-95%. The error model excluded 1,161 of the 2,366 raw unfixed substitution calls (49%, **Supplementary Fig. 2e-f**) in the set of samples without personal genomes, suggesting that a large proportion of raw unfixed variant calls in short-read data are false calls. We similarly excluded insertion and deletion (indel) calls predicted to be false, retaining 278 unfixed indels (**Methods**, **Supplementary Results**). Across the 1,878 unfixed substitutions and indels, the median within-host frequency is 15% (IQR = 30%).

At baseline, there was a median of 1 unfixed variant in each host (range 0-20) with no significant differences by lineage (p = 0.69, likelihood ratio test across lineages 2, 3, and 4). The measured number of unfixed variants within-host suggests that the sputum culture and sequencing bottleneck is <0.1% of the bacilli sampled from sputum and <0.001% of the bacterial population size estimated to exist in human cavitary TB disease (estimated at >10^5^ bacilli, and accounting for *Mtb’s* known slow mutation rate; **Methods**).^27^

### Unfixed variants in drug resistance genes are rare in rifampicin-susceptible TB

Unfixed variants in genes known to be involved in drug resistance are rare among clonal infections in this cohort. Only 7 participants (2%) with clonal infections had non-silent variants in and around genes known to cause drug resistance in *Mtb* (**Table 2**). One of the participants was infected with a phenotypically resistant isolate to isoniazid (INH) and demonstrated four low frequency variants (frequency 7-31%) in *katG*. Another participant had an INH-susceptible infection and demonstrated one unfixed variant at a frequency of 11% in *katG*. All five variants were not detected on follow-up sequencing. Five other participants had unfixed variants in *gyrB, fbiA, fbiC,* and *pncA*. These genes are known to encode fluoroquinolone, delamanid, and pyrazinamide resistance, but the specific variants are not described in the WHO drug resistance catalog.^2^

**Table 2.** Variants in and around known antibiotic resistance genes in the TRUST cohort. Of the variants, only two are in the 2023 WHO drug resistance mutation catalog^2^ – katG_p.Trp149Arg and katG_p.Tyr98Cys, which are both associated with isoniazid resistance by the relaxed thresholds.^2^ Only participant T0331 had any phenotypically measured first-line drug resistance, which was to isoniazid.

| Participant ID | Position in H37Rv Coordinates | Variant | AF Change | Isoniazid Resistant |
| --- | --- | --- | --- | --- |
| T0058 | 6,923 | gyrB_p.Phe562Val | 7.1% → 11% | No |
| T0069 | 2,155,174 | katG_p.Ile313Thr | 11% → 0% | No |
| T0106 | 3,640,843 | fbiA_p.Trp101Arg | 76% → 0% | No |
| T0210 | 6,078 | gyrB_p.Gly280Val | 0% → 9% | No |
| T0310 | 2,288,760 | pncA_p.Ala161Val | 5% → 0% | No |
| T0331 | 2,153,995 | katG_p.Ala706Glu | 31% → 0% | Yes |
|  | 2,154,800 | katG_p.Trp438Arg | 26% → 0% |  |
|  | 2,155,667 | katG_p.Trp149Arg | 11% → 0% |  |
|  | 2,155,819 | katG_p.Tyr98Cys | 7% → 0% |  |
| T0453 | 1,303,813 | fbiC_p.Phe295Leu | 0% → 6% | No |

Among the 25 participants with mixed infections, 19 (76%) have unfixed non-silent variants in drug resistance genes (**Supplementary Table 2**).^2^ Mixed infection samples are more likely than clonal samples to have non-silent unfixed variants in drug resistance genes (p = 1.2 x 10^-21^, one-sided Fisher’s exact test). Unfixed variant burden is stable longitudinally, but some variants rise to fixation

Among the 284 participants with clonal infections and longitudinal sampling, the unfixed variant count is stable between baseline and follow-up (median difference 0, mean difference 0.28 variants, p = 0.15, two-sided paired t-test, **Fig. 1c**), and the two values are highly correlated (Spearman ρ = 0.42, p = 1.2 x 10^-13^). Stratifying by allele fraction (**Table 3**), we observe that the largest differences in unfixed variant count between baseline and follow-up occur at low allele frequencies, suggesting that more rare variants (AF ≤ 10%) have higher turnover.

**Table 3.** Changes in the number of unfixed variants for 284 participants between baseline (BL) and follow-up (FU) sequencing. The median and mean differences are follow-up counts minus the baseline counts. All allele frequency ranges are bound-inclusive.

| Variant AF Range | Mean (Median) Difference, | Two-sided paired t-test |
| --- | --- | --- |

|  | Follow-Up – Baseline | p-value |
| --- | --- | --- |
| [5%, 10%] | 0.15 (0) | 0.16 |
| [10%, 25%] | 0.10 (0) | 0.26 |
| [25%, 95%] | 0.02 (0) | 0.84 |

Of the 1,269 unfixed variants at either timepoint, the majority (1,067, 84%) are detected at only one time point; 140 (11%) are unfixed at both time points, and the remaining 62 (5%) rise to or decrease from fixation. Across these scenarios, the average allele frequency increases over time (average ΔAF = 0.11, p = 6.8 x 10^-36^, one-sided paired t-test), but this average change is driven by a small subset of variants with large increases in frequency (n=154, ΔAF > 0.5, **Fig. 1d**). Overall, the observed temporal dynamics demonstrate a high turnover of unfixed variants and/or a stochasticity of sampling through sputum culture from a relatively stable sized population over the sampling time (∼6 weeks). Despite this high flux and/or stochasticity, within-host sequencing is able to identify a subset of variants that increases in frequency over time and may therefore be adaptive to the pathogen.

### Within-host diversity is enriched in genes mediating pathogen adaptation and drug tolerance

To determine whether within-host genetic diversity is driven by active pathogen adaptation rather than neutral random changes, we assessed the distribution of unfixed variants across the *Mtb* genome (**Methods**). Across the cohort, we identified 781 unfixed single nucleotide variants (SNVs) at baseline (in n=261 participants; 134 participants had no unfixed variants) and 651 at follow-up (n=206 participants). Rather than occurring randomly, non-silent unfixed SNVs were significantly enriched in several gene functional groups, including transcription factors^28–31^, transporter proteins,^32,33^ and two component systems^34–36^ (**Fig. 2a**, **Table 4, Supplementary Data 6**). Phylogenetic analysis revealed that many of these variants emerged independently across the four lineages in our cohort, indicating convergent evolution in response to a shared host related selective force **(Fig. 3)**.^37^ The most frequent unfixed variant across the cohort was a premature stop codon in *sugI*, a putative sugar transporter, which arose independently in 17 patients across four lineages. Among the unfixed variants that increase by more than 50% in frequency over time, the most common occur in *sugI, phoT, devR,* and *phoR*. Hence, the genes enriched in unfixed variants at baseline are also most likely to have variants that increase in frequency over time, supporting a potential adaptive role for these variants within the host under treatment pressure.

**Figure 2.**
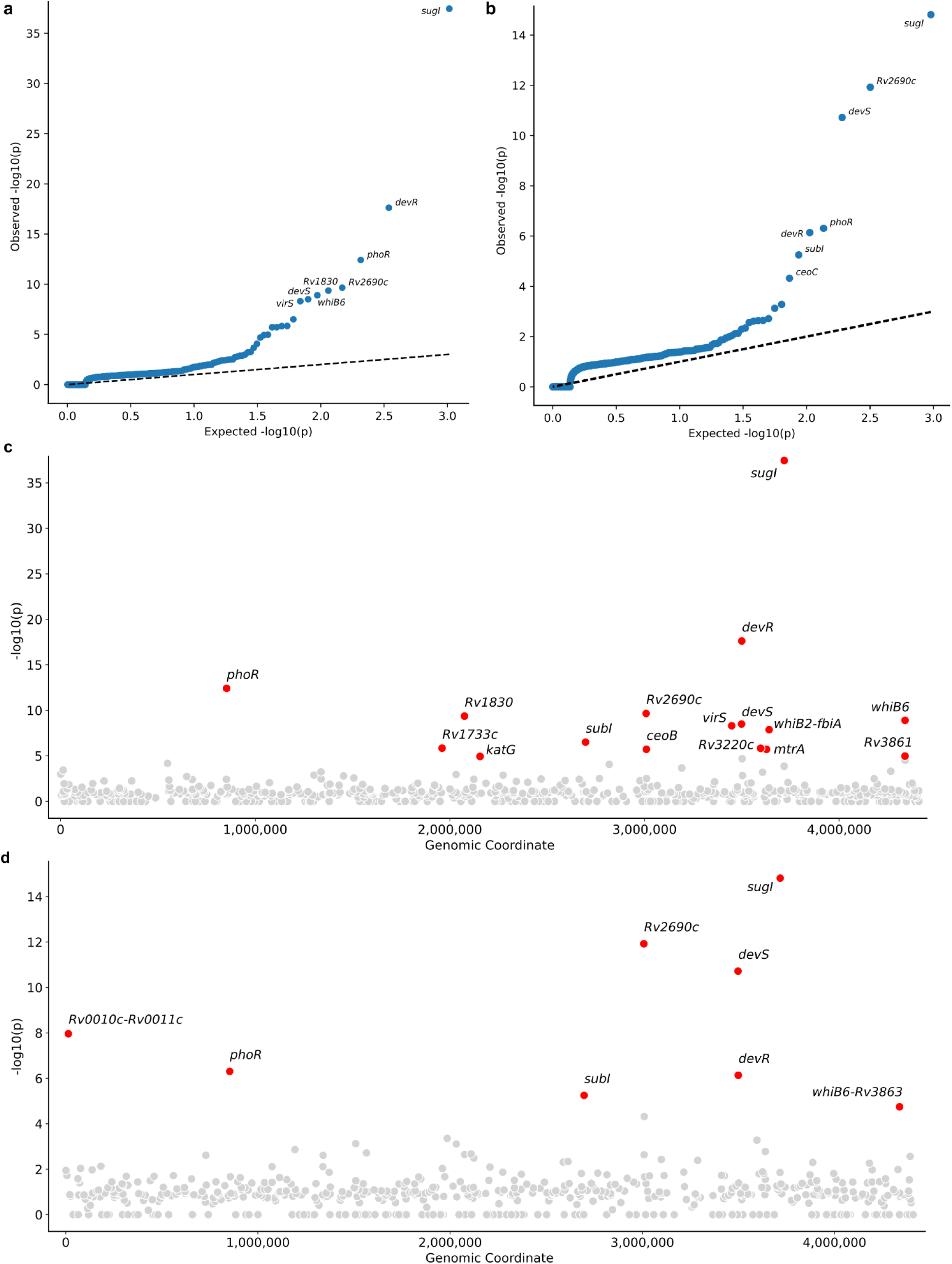
Gene-level enrichment of non-silent unfixed SNVs. **a-b**: QQ plots of one-sided Poisson test p-values for SNVs at baseline (**a**) and follow-up (**b**). The most highly enriched genes or intergenic regions are labeled by name. **c-d**: Manhattan plots of the negative logarithm of raw Poisson p-values for baseline (**c**) and follow-up (**d**). Genes with a Bonferroni-corrected p-value < 0.01 are colored in red and labeled, and all other genes are colored in gray. Coordinates are in H37Rv in panels **c** and **d**. Unfixed SNVs were found in all four lineages, but there is only a single SNV in a single L1 sample taken at baseline and no L1 samples at follow-up.

**Figure 3.**
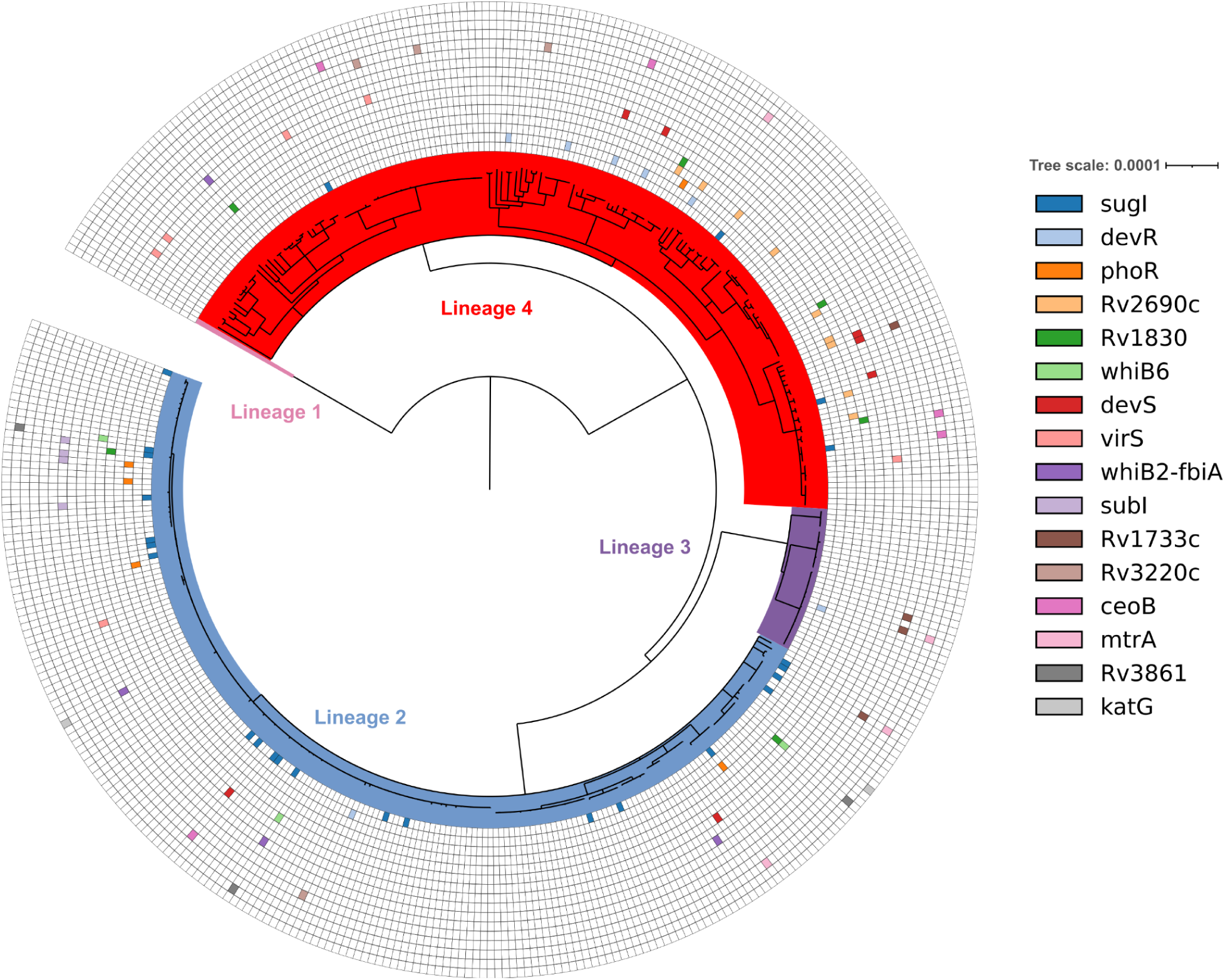
Maximum likelihood tree of 395 baseline whole-genome sequencing samples. The tree is annotated with 16 concentric circles for the 16 regions enriched in unfixed SNVs in **Table 4**. A colored square indicates that a sample contains an unfixed SNV (a variant with AF ≥ 0.05 and AF ≤ 0.95) in the specified region. The regions are ordered by increasing p-value, as in **Table 4**. The four lineages represented in the dataset are labeled by color and with text.

**Table 4.** Sixteen regions with non-silent unfixed SNVs with Bonferroni-corrected enrichment p-values < 0.01 at baseline. . Intergenic variants are listed with the two flanking genes separated by a hyphen. The *whiB2-fbiA* variant is upstream of both genes, and this region contains a known *resR* binding site.^29^ “Count” is the number of participants in which the unfixed SNVs in each region occurred, but the SNVs are not identical across participants. “Lineages” is a comma-separated list of the lineages in which the unfixed SNVs occur. For the intergenic regions, the “Function” column reflects only the downstream genes. If both genes are downstream of the variant, the functions are listed in the order in which they appear, separated by a comma. Regions are ordered by increasing p-value. Full enrichment results are in **Supplementary Data 6.**

| Region | Function | Count | Bonferroni p-value | Lineages |
| --- | --- | --- | --- | --- |
| *sugI* | Sugar transporter | 25 | $1.8 \times 10^{-35}$ | 2, 4 |
| *dosR* | Two component response regulator | 7 | $1.2 \times 10^{-15}$ | 2, 3, 4 |
| *phoR* | Two component sensor kinase | 5 | $2.0 \times 10^{-10}$ | 2, 4 |
| *Rv2690c* | Transporter protein | 9 | $1.1 \times 10^{-7}$ | 4 |
| *Rv1830 (resR)* | Transcription factor | 6 | $2.2 \times 10^{-7}$ | 2, 4 |
| *whiB6* | Transcription factor | 3 | $6.5 \times 10^{-7}$ | 2 |
| *dosS* | Two component sensor kinase | 7 | $1.6 \times 10^{-6}$ | 2, 4 |
| *virS* | Transcriptional regulator of virulence genes, including the *mymA* operon | 6 | $2.5 \times 10^{-6}$ | 2, 4 |
| *whiB2-fbiA* | Transcription factor, Coenzyme F420 production | 4 | $7.9 \times 10^{-6}$ | 2, 4 |
| *subI* | Sulfate transport | 4 | $1.6 \times 10^{-4}$ | 2 |
| *Rv1733c* | Latent TB-associated antigen, part of the *dosR* regulon<sup>40,41</sup> | 4 | $7.4 \times 10^{-4}$ | 2, 3, 4 |
| *Rv3220c* | Two component sensor kinase | 4 | $7.7 \times 10^{-4}$ | 2, 4 |
| *ceoB* | Potassium transporter | 5 | $9.9 \times 10^{-4}$ | 2, 4 |
| *mtrA* | Two component response regulator | 4 | $1.0 \times 10^{-3}$ | 2, 3, 4 |
| *Rv3861* | Unknown | 3 | $5.5 \times 10^{-3}$ | 2 |
| *katG* | Activates isoniazid | 2 | $6.0 \times 10^{-3}$ | 2 |

In addition to SNVs, we studied phase variants - rapid, reversible, short insertions or deletions occurring in hypermutable repetitive DNA sequences (homopolymeric tracts - **Methods**). Phase variation occurs at a faster rate than SNVs due to replication error^38^ and is reported to affect drug tolerance^38,39^ and virulence in *Mtb*.^19^ Of 278 indels identified, 28 (10%) were phase variants. The majority (16 of 28) occurred in genomic in regions known to be associated with drug resistance and/or virulence,^38^ including variants that regulate *espR* and increase expression of *espA* a canonical *Mtb* virulence factor.^19^ Although rare (n=2), the observed *espR* phase variants rose to fixation in both participants during treatment (**Supplementary Table 5**). Together, these patterns suggest that within-host diversity includes adaptive remodeling of pathways important for *Mtb* survival during treatment.

### Baseline within-host diversity predicts unfavorable treatment outcomes

To evaluate the prognostic value of within-host pathogen diversity, we analyzed data from 364 participants who had whole genome sequencing performed within the first two weeks of treatment initiation. The participants had time-to-culture-conversion (TCC) assessed from weekly cultures during the first 12 weeks on treatment and at 5 months, as well as composite unfavorable outcome (treatment failure, death, and relapse) assessed over a median of 1.5 years (IQR = 0.13 years) of post-treatment follow up. All participants were monitored 5 days a week for treatment adherence. 25 individuals (7%) had an unfavorable outcome: 10 treatment failures, 3 deaths, and 12 relapses.

Baseline clonal within-host diversity (*i.e.,* unfixed variant burden) is not associated with cavitation (p = 0.75, two-sided Mann-Whitney U test), smear positivity (p = 0.97, two-sided Mann-Whitney U test), percent of lung involved with TB (PLI, Spearman ρ = 0.09, p = 0.08) or time to positivity of culture (TTP, Spearman ρ = 0.06, p = 0.28), and is not significantly different across the 5 smear grade levels (likelihood ratio test p = 0.79).

Although at least one prior study has reported mixed lineage infections to be associated with unfavorable treatment outcomes,^42^ in this cohort, a mixed lineage infection was detected in only 5% of sequenced baseline cultures. These mixed lineage infections were not associated with unfavorable outcomes (**Supplementary Results**). Below, we exclude participants with mixed lineage infections to isolate the effect of within-strain (*i.e.,* clonal) evolutionary diversity on outcome.

In Kaplan Meier analysis, baseline within-host diversity >3 unfixed variants is associated with time to composite unfavorable outcome (log-rank test p = 0.04, **Methods, Supplementary Fig. 5a**) but is not associated with TCC (log-rank test p = 0.22, **Supplementary Results**). This suggests that unfixed variant burden is more predictive of delayed events like relapse than earlier metrics of culture conversion. In line with this, we observed that survival differed beyond 12 months from treatment initiation between participants with >3 unfixed variants (vs. ≤3) but not within 12 months (log-rank test p = 0.14 vs. 0.04 for time ≤ 12 months *vs.* the full observation time respectively).

To confirm that pathogen diversity is predictive of composite unfavorable outcomes independent of known clinical risk factors, we built a multivariate Cox proportional hazards survival model adjusting for demographic and clinical covariates: age, sex, BMI, smoking status, HIV/CD4+ T cell count, diabetes, prior TB disease, smear positivity, and percent lung involved (PLI). In the multivariate model, severe immune suppression (HIV positivity with CD4 < 200 cells/mm^3^) and PLI are associated with composite unfavorable outcomes (p-values < 0.05, **Fig 4d**). TTP at baseline was removed from the multivariate model because it was missing for 13 participants and did not demonstrate a univariate association with outcomes (p = 0.72, **Fig. 4c**).

**Figure 4.**
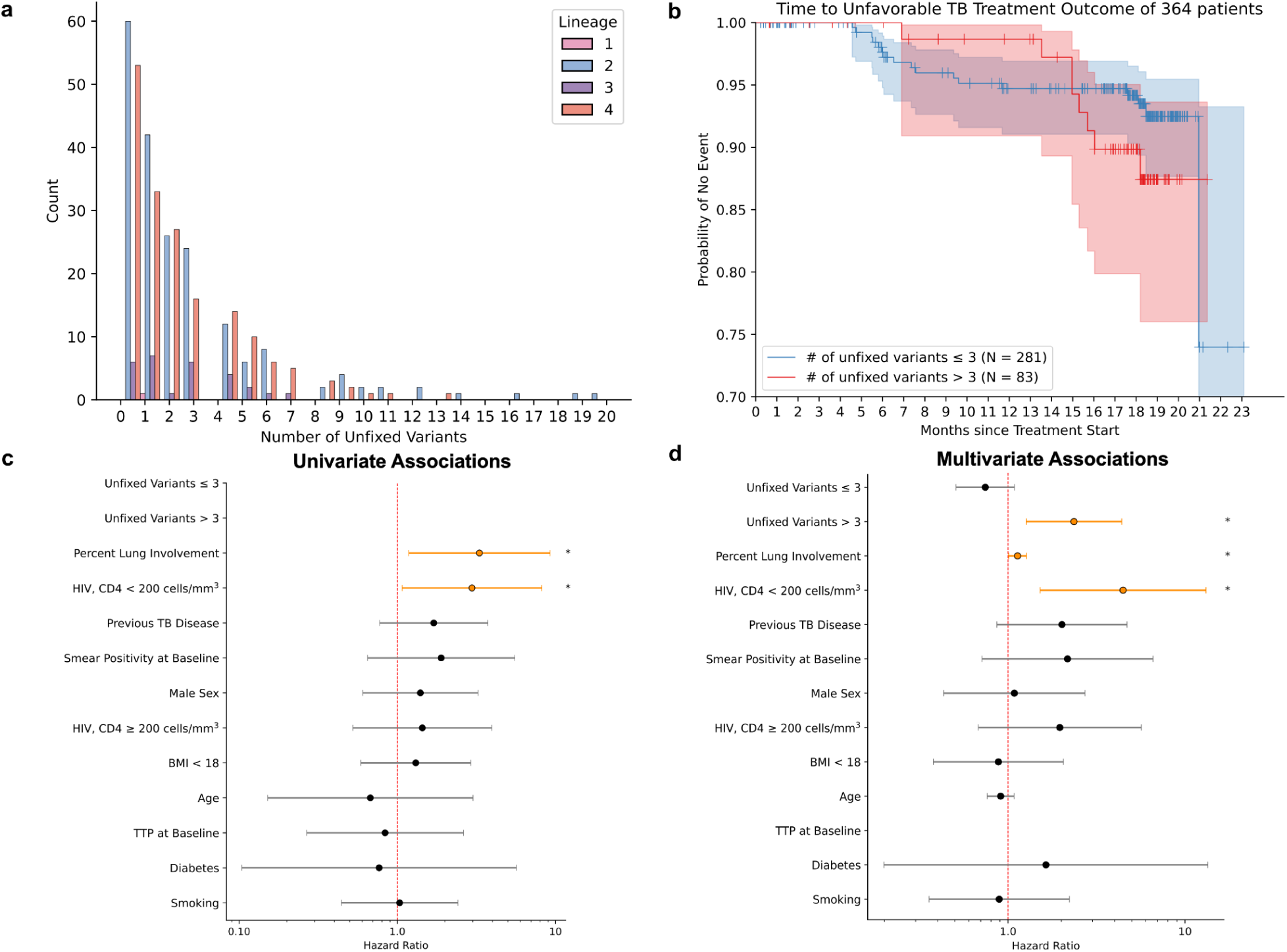
Unfixed variant burden at baseline is associated with unfavorable outcomes. a: Distribution of number of unfixed variants (substitutions and indels) per sample, colored by primary lineage, across 395 baseline samples. **b:** Kaplan-Meier curves stratified by the number of unfixed variants at baseline. The y-axis extends from 0.7 to 1 to better visualize the differences between curves. **c:** Forest plot of univariate associations in Cox proportional hazards model between patient covariates and outcomes. Unfixed variant burden was not tested for univariate association with outcomes. **d**: Forest plot of multivariate associations between unfixed variant burden and outcomes, adjusted for the covariates in **c**, except for TTP at baseline, which is missing for 13 participants and was not associated in the univariate model (p = 0.72). The forest plots in panels **c** and **d** contain hazard ratios with 95% confidence intervals (Wald). Standard errors were computed by the Cox regression fitter in lifelines. Variables with two-sided p-values < 0.05 are shown in orange and denoted by an asterisk. The hazard ratios for age, unfixed variant burden > 3, and percent lung involvement (PLI) are per 5 unit increases. For individuals with pathogen diversity ≤ 3 unfixed variants, baseline diversity does not predict composite unfavorable outcomes.

For individuals with pathogen diversity of >3 unfixed variants at baseline, diversity as a continuous predictor is significantly associated with treatment outcome. For these individuals, every increase of 5 unfixed variants was associated with more than a doubling of the risk for an unfavorable outcome (adjusted Hazard Ratio [aHR], 2.36, 95% CI 1.27–4.39; p = 0.007, **Fig. 4d**). Across an unfixed variant count of 0-3, we observed no relationship between diversity and unfavorable outcome (**Fig. 4d**). Having >3 unfixed variants is positively associated with having an unfixed variant in the 16 regions found to be enriched in unfixed variants in this cohort (**Fig. 3**) (p = 2.8 x 10^-10^, χ^2^ test). These individuals with >3 unfixed variants are also more likely to have phase variants (p = 8.8 x 10^-4^, one-sided Fisher’s exact test).

We tested if pathogen diversity measured on follow up was more predictive of unfavorable outcomes than baseline diversity as, hypothetically, follow up diversity may more directly assess treatment response. 262/364 participants were included in this analysis because the remaining individuals did not have clonal infections with a longitudinal follow-up sequence available. Unfixed variant burden at follow up (median of 6 weeks from baseline) had a similar association with unfavorable outcome in the adjusted Cox model (aHR = 2.26, 95% CI = 1.11–4.58, p = 0.02, **Supplementary Results**). However, the Cox model with follow up diversity has lower model fit compared with the Cox model with baseline diversity (AIC 216.2 vs 212.4 respectively on the same participant subset). This observation supports sampling at baseline as the preferred timepoint, and provides evidence for robustness of the association to sampling time within the first two months of therapy.

## Discussion

In this prospective cohort study, we demonstrate that within-host genetic diversity of *M. tuberculosis* at the time of diagnosis is a strong independent predictor of unfavorable treatment outcomes. By leveraging long-read personal reference genomes to minimize bioinformatic error, we found that unfixed variation in clonal infections is relatively rare, reflecting *Mtb* low evolutionary rate and a substantial sampling and culturing bottleneck. However, when present, this baseline variant burden strongly associates with unfavorable post-treatment outcomes, including relapse. These findings highlight the relevance of within-host heterogeneity and its potential role as a novel prognostic biomarker to guide duration of treatment.

The independent predictive power of within-host diversity suggests that genomic heterogeneity captures a new dimension of disease severity missed by standard assessments. We hypothesize that the total burden of within-host variants may act as a molecular proxy for the effective population size of infection and hence a measure of viable bacterial burden of infection. Traditional metrics, such as smear grade on microscopy, or cycle threshold available through nucleic acid amplification tests, are inherently limited by their inability to distinguish live, actively replicating bacilli from dead organisms. Metrics like time-to-positivity in culture, and interval culture are limited by the culture bottleneck itself and their inability to capture viable but non-culturable cells.^43^ Radiographic cavitary and percent of lung involvement reflects cumulative host tissue damage and the host immune state rather than real-time pathogen viability. In contrast, principles of neutral population genetics dictate that the accumulation of genetic diversity requires sustained, active bacterial replication over time. Therefore, a more diverse within-host population suggests a more chronic, unconstrained infection that has reached a larger viable bacterial burden and that this associates with a higher risk of relapse and other unfavorable post-treatment outcomes.

In addition to reflecting global dynamics of the infecting bacterial population, within-host diversity also offers the pathogen greater opportunity for adaptation to drug or other within-host pressures. We observed significant enrichment of unfixed variants in transcription factors known to affect virulence^30,44^ and *Mtb* growth after antibiotic exposure, including *resR*.^29,45^ Unfixed variants were also enriched in genes involved in drug resistance, drug tolerance, and host pathogen interactions. Specifically, we find enrichment in transporter proteins and two-component regulatory systems, including the *dosRS* regulon, which governs mycobacterial virulence and entry into a persistent drug tolerant state under host stress.^36,46^ The concentration of variants in these specific pathways suggests that they may be on the causal pathway linking overall diversity and unfavorable treatment outcomes, *i.e.,* by allowing subpopulations of bacteria to survive multi-drug therapy. This persistence phenotype also aligns with the temporality of the clinical association, explaining why baseline diversity predicts delayed clinical outcomes like relapse rather than the earlier treatment metric of time-to-culture conversion.

A major strength of this study is the rigorous methodological framework used to call unfixed variants such that they capture true genetic diversity rather than bioinformatic error. Historically, accurately calling minority variants from short-read data has been severely compromised by reference bias, leading to overcalling minority variants or requiring a high degree of manual curation. By building high quality personal hybrid assemblies using long-read data and training a model to predict short-read variant accuracy, we were able to exclude the majority of artifactual variant calls. This model can be incorporated in the bioinformatic workflows of others analyzing lineage 2, 3 and 4 genomes to improve the accuracy of unfixed variant calls.^26^ Ultimately, our results demonstrate that while true within-host variant burden is lower than some prior estimates suggest,^11,26,29^ it is clinically and biologically relevant.

Our study has several limitations. First, the cohort reflects the local TB epidemiology in South Africa, where disease burden may be higher than other settings, and the *Mtb* lineage distribution consists primarily of lineages 2, 3, and 4. Validating the variant calling pipeline and confirming the optimal prognostic threshold of variant burden will require application to more diverse, globally representative clinical cohorts. Second, sequencing was performed on cultured isolates rather than directly from sputum. While the impact of culture on within-host diversity and the size of this bottleneck have been previously debated, our data supports that this bottleneck is substantial. The bottleneck likely results from differential *in vitro* growth rates and/or stochastic attrition.^47–49^ In addition, others have shown that when within-host diversity exists, it is often distributed across different lesions (granulomas),^13,50,51^ suggesting that there is likely stochasticity in capturing the full within-host diversity in expectorated sputum. In the future, direct from sputum sequencing combined with pooling of multiple sputum samples may allow for future refinement of this biomarker. Given the cost, biohazard, and delays incurred by *M. tuberculosis* culture, an ideal metric for predicting TB treatment response would need to be measurable rapidly and directly on the patient sample.

In conclusion, we provide a robust framework for identifying true unfixed genetic variation in *M. tuberculosis*. As targeted and whole genome sequencing become increasingly integrated into global TB diagnostic labs for resistance testing, evaluating within-host diversity will become increasingly accessible to extract additional prognostic information. Assessing pathogen diversity at baseline is a promising new tool for stratifying patient risk, with potential implications for guiding the use of novel short-course regimens.

## Code Availability

All original figures and the code to perform the analyses and generate figures are available at https://github.com/sanju99/MtbLongitudinalDiversity.

## Data Availability

All data produced are available online at https://github.com/sanju99/MtbLongitudinalDiversity
Sequencing data will be deposited in the Sequence Read Archive (SRA), and public identifiers will be updated in the GitHub repository upon creation.

## Acknowledgements

We thank the study participants and study staff for making this work possible. Computational resources and support were provided by the Orchestra High Performance Compute Cluster at Harvard Medical School, which is funded by the NIH (NCRR 1S10RR028832-01). S.G.K. was supported by the National Science Foundation Graduate Research Fellowship DGE2140743. M.R.F. was supported by a National Institute of Allergy and Infectious Diseases / National Institutes of Health grant R01AI155765. R.M.W. was supported by the South African Medical Research Council.

## Methods

### A. Short read variant calling against the H37Rv reference genome

Illumina reads were preprocessed using fastp (v1.0.1) for adapter trimming and removal of reads shorter than 50 base pairs, and only reads mapped to the *Mycobacterium tuberculosis* complex (taxid 77643) and its parent and child taxids by kraken2 (v2.1.3) using the standard database (downloaded in June 2020) were retained. These processed Illumina reads were subsequently used for variant calling against the personal reference genomes and the H37Rv reference genome.

These processed Illumina reads were aligned to the H37Rv reference genome (NCBI RefSeq NC_000962.3) using bwa mem (v0.7.19) with a seed length of 80. We then marked duplicates with picard (v3.4.0) and performed variant calling with freebayes (v1.3.10) using a minimum mapping quality of 30, a minimum base quality of 30, a minimum alternate allele count of 2, and a minimum allele fraction of 0.01. All haplotypes were returned and later separated.

### B. Hybrid genome assembly and variant calling

PacBio HiFi reads were assembled using 3 iterations of flye (v2.9.2), circularized with circlator (v1.5.5), and then polished with the processed Illumina reads in **Methods Section A** using pilon (v1.23) to generate personal reference genomes. Gene annotations were lifted from H37Rv to the personal genomes using liftoff (v1.6.3). To call unfixed variants from the personal reference genomes, we aligned Illumina reads from the previous step to the personal genome using bwa mem (v0.7.19) with a seed length of 80. We then marked duplicates with picard (v3.4.0) and performed variant calling with freebayes (v1.3.10) using a minimum mapping quality of 30, a minimum base quality of 30, and a minimum allele fraction of 0.01. Unfixed variants were transferred from the personal genome coordinates to H37Rv coordinates using paftools liftover, which is part of the minimap2 suite (v2.30).

### C. Decomposing complex variants into individual variants

Because freebayes is a haplotype variant caller, multiple variants supported by the same reads are listed as a single haplotype in the output variant call format file. To estimate the number of individual SNVs, complex variants, where an indel and an SNV occur on the same haplotype, needed to be split. We did this using vcfwave to decompose complex variants, followed by bcftools split multiallelics to split sites with more than two alleles.

However, at multiallelic sites, vcfwave and bcftools do not split the read counts supporting different alternative alleles properly, so we wrote a custom Python script to count the number of reads supporting each alternate allele by matching cigar strings. Multi-nucleotide substitutions were easily split into the component SNVs using custom Python code.

### D. Logistic regression model to predict the accuracy of an unfixed SNV

The logistic model was only run on variants with 0.05 ≤ AF ≤ 0.95. The included features were the number of discordantly paired reads normalized to coverage, average base quality of bases supporting the variant, ratio of coverage at the site to the rolling average, the absolute value of the difference between 0.5 and the proportion of reads supporting the variant that are in the forward orientation, number of soft clipped bases normalized to coverage, number of soft clipped reads supporting the variant normalized to total reads supporting the variant, and the median of the normalized intra-read position at which the variant occurs.

The coverage rolling average was computed with a window size of 100 base pairs, and we took the maximum of the rolling averages computed from the up-and downstream directions. We split the coverage variable into two variables to fit individual coefficients for coverage increases and decreases. Given a coverage ratio of *r_c_*, when *r_c_* ≥ 1, the coverage increase variable is equal to *r_c_*-1 and 0 otherwise. Conversely, the coverage decrease variable is equal to 1-*r_c_* when *r_c_* <1 and 0 otherwise.

The model was trained on 802 candidate unfixed SNVs (401 real, 401 not real) from the 172 WGS samples with matched personal reference genomes. The variants themselves were derived from variant calling against H37Rv, and the real labels were derived from comparing to the unfixed SNVs from variant calling with the personal reference genomes. After fitting the model, the classification threshold for dichotomizing predicted probabilities was selected to be 0.67 to maximize the F1 score, resulting in 9 false positives and 9 false negatives.

### E. Adjusting allele fractions to correct false unfixed indels

Calling unfixed indels is generally less affected by reference bias because variant callers require greater evidence to call indels than SNVs. The primary issue in accurately calling low frequency indels is reads not sufficiently covering an indel. These reads artificially bring down the allele fraction of the indel, making it appear as though a fixed indel is unfixed and inflating the number of unfixed indels when simply thresholding on allele fraction.

Indels were called using the same pipeline for substitutions and the same freebayes parameters as in **Methods Section A**, followed by left-alignment and normalization with bcftools (v1.21). For each indel with 0.05 ≤ AF ≤ 0.95, we extracted all the reads from the pileup at the position where the indel begins. We then excluded all reads with soft clipping and all reads that start or end within 10 base pairs of the putative indel, including reads that terminate in the middle of the indel. We then determined if the start and end positions for reads that support the indel are significantly different from those of reads that do not support any indel. We compared the medians of the distributions using two Mann-Whitney U tests, one for the start positions and one for the end positions. If both tests returned p-values < 0.01, then this was considered evidence that the reads that do not support the indel do not sufficiently cover it. These reads would be unable to support an indel, even if it truly exists, and so they too were excluded.

We then recomputed the allele fraction using all remaining reads that do and do not support the indel (if both p-values from the Mann-Whitney U tests were < 0.01, then these two values are equivalent, and the new AF is 1). Indels at sites with a coverage less than one-third of the genome-wide median were also excluded. For substitutions, we used a threshold of one-half of the genome-wide median, but this was found to be too strict for indels. Finally, indels occurring within transposable elements, phage sequences, and ribosomal RNA regions were excluded, as was done for substitutions.

### F. Estimating the sputum, culture and sequencing bottlenecks

We estimated the number of bacterial cells sampled and sequenced relative to the total bacterial population in the lung under a simplified population genetic framework for a within-host *Mtb* population. We assume an infinite-sites neutrally evolving haploid population with an effective population size (*N_e_*) of 10^5^ corresponding to the lower end of estimates proposed for cavitary disease. Within host *Mtb* population sizes have been hypothesized to range from 10^5^ to 10^8^.^27^

The mutation rate (μ) was assumed to be 1.4 x 10^-3^ SNPs per genome per generation^38,52^. This value was derived from the reported estimate of approximately 0.5 SNPs per genome per year, scaled to a 24-hour *Mtb* generation time by dividing by 365 days per year.^52^ The expected number of segregating sites E(*S*) corresponds to the number of unfixed variants within host, and the relationship between this variable, and other parameters is given by:

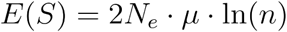

where *n* is the number of bacterial cells sampled and sequenced relative to the total bacterial population in the lung. This approximation assumes that *n* << *N_e_*, which was considered reasonable under the present sampling framework.

We solved for *n*, the effective sequencing sample size, using the observed range on the number of unfixed variants per population (1 to 19). The calculation estimates *n* at ∼1, and a overall bottleneck of <0.001% for an *N_e_* of >10^5^. Given that sputum samples for smear positive/cavitary TB are expected to carry >10^3^ bacilli in the volume inoculated into culture (a bottleneck of 1% from the 10^5^ bacilli in the lung), the culture and sequencing bottleneck itself is estimated at an additional 0.1% of the bacilli sampled from sputum.

### G. Structural variant detection

Structural variant detection was performed using long-and short-reads separately. For the former, PacBio HiFi reads were aligned to the H37Rv reference genome using minimap2 (v2.30), followed by structural variant calling with sniffles (v2.7.1) with default settings, and finally left-alignment and normalization with bcftools (v1.21). We excluded variants that failed to meet the minimum read support filter indicated by sniffles. For short reads, we ran delly (v1.7.2) with default settings on the BAM files generated in **Methods Section A**. We included only variants that had at least 10 paired-end reads of support.

### H. IS*6110* element detection

To detect IS6110 elements, the 1,355-base pair long IS*6110* element sequence was extracted from the H37Rv genome and aligned to each of the 172 hybrid assemblies using minimap2 (v2.30). The alignment coordinates were then transferred to H37Rv coordinates using paftools liftover to be able to compare the locations of the IS*6110* elements in each hybrid assembly in a standardized coordinate system. The number of IS*6110* elements ranged from 1-25 per sample, and the average numbers per lineage are 1 for lineage 1, 22.5 for lineage 2, 16.7 for lineage 3, and 9 for lineage 4.

### I. Poisson test for gene-level unfixed SNV enrichment

We assumed that the number of unfixed single nucleotide variants (SNVs) in each gene or intergenic region follows a Poisson distribution parameterized by the empirical unfixed mutation rates. These rates were computed for each of the 12 types of single base pair substitutions, ignoring surrounding context, separately at baseline and follow-up. As expected, the four transition mutations have the largest rates (**Supplementary Table 4**).

Gene and intergenic region bounds were extracted from the H37Rv genome coordinates.^53^ We estimated the genome-wide unfixed substitution mutation rate, λ(*r*→*a*) for a given substitution *r*→*a*, where *r* is the reference nucleotide, *a* is the alternative nucleotide, *r,a* ɛ {*A,C,G,T*}, and *r* ≠ *a*, ignoring the surrounding context.

For a given gene *g* with codons in the set *S_g_*, the expected number of unfixed SNVs is

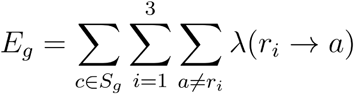

where *c*=(*b*_1_,*b*_2_,*b*_3_) is each codon, *r_i_* is the reference (H37Rv) nucleotide at position *i* in codon *c*, and *a* is the alternative nucleotide. For a given intergenic region *g* with length *L_g_*, the expected number of unfixed SNVs reduces to

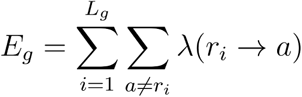

The expected number of unfixed variants per region therefore accounts for both the region length, nucleotide composition, and for coding regions, the sequence of the codon in which the variant occurs. For coding regions, we tested for enrichment of non-silent variants, and for intergenic regions, we tested for enrichment of all variants. Enrichment was determined by comparing the number of observed unfixed SNVs for each group to the expected value using a one-sided Poisson test.

### J. Phylogenetic tree construction

To extract SNVs with which to build a phylogenetic tree, we ran pilon (v1.24) on the BAM files generated in **Methods Section A** with a minimum mapping quality of 1 to exclude multiply mapped reads. We used pilon because it is not a haplotype variant caller, so it is more straightforward to extract individual SNVs. We extracted SNVs with allele frequency > 95%, mapping quality ≥ 40, base quality ≥ 30, and read depth ≥ 10.

We kept only baseline (within the first two weeks) WGS samples with F2 ≤ 0.03 (N = 395). We excluded any position with an overlapping deletion or low-quality SNV anywhere in the dataset, leaving 14,049 SNVs with which to build the tree. A maximum likelihood tree was built using iqtree (v2.4.0) with ascertainment bias correction, searching for the best fitting nucleotide substitution model, and allowing multifurcations due to short branch lengths. The tree was visualized in FigTree, and the single lineage 1 sample in the dataset was used as the outgroup. Branch support was assessed using the approximate Bayes test.^54^

### K. Clinical treatment outcomes models

To test if the number of unfixed SNVs at baseline is associated with unfavorable TB treatment outcome, we built multivariate Cox proportional hazards models to associate clinical and microbiological data from participants in the TRUST study^21^ with two events – sputum culture conversion and unfavorable outcomes. Participants in this study have rifampicin-susceptible pulmonary TB diagnosed by GeneXpert and were all treated with a 6-month regimen of RIPE (rifampicin, isoniazid, pyrazinamide, and ethambutol), followed by 12 months of follow-up. Time zero for each participant is the time of initial screening and treatment initiation.

A total of 379 participants remained after excluding participants who changed treatment regimens due to developing rifampicin resistance or needing a liver-friendly regimen, participants without WGS of their bacterial cultures performed within the first two weeks of enrollment, and participants with incomplete data. We adjusted the Cox models using 9 relevant clinical confounders that were selected based on past association with poor TB treatment outcomes.^55–57^ Percent lung involvement was read by human physicians as done previously.^58^ The other independent variables were determined from clinical records. Restricting to participants with clonal *Mtb* infections with F2 ≤ 0.03 and no missingness among the 9 confounding variables, we retained 364 participants for the outcomes models.

Unfavorable treatment outcomes were defined as treatment failure, TB disease relapse, or death. Treatment failure was assessed after 6 months of treatment, either by a positive culture or symptomatically. Relapse was assigned if a participant was sputum-or smear-positive at the conclusion of the 12-month follow-up period. The median duration of enrollment of the 364 individuals was 546 days, and the longest was 693 days. Follow-up periods were longer than 12 months for some individuals due to appointment delays during the COVID-19 pandemic. We censored participants whose deaths were attributed to non-TB causes: cancer (N = 3), cardiovascular disease (N = 1), and accidental deaths (N = 1). All other patients were right-censored at their study completion date or their last known clinic appointment if they were lost to follow-up (N = 31) or still enrolled in the study (N = 11) at the time of analysis.

Sputum was sampled weekly for the first 12 weeks of treatment. Time to negative sputum culture conversion (TCC) was defined as the number of weeks between the time of treatment initiation and the first of two consecutive *Mtb*-negative sputum cultures not followed by another positive culture. To be included in the TCC analysis, participants had to have at least 3 uncontaminated sputum samples and a positive *Mtb* culture within the first 5 weeks of initial screening. We imputed missing cultures after this using the mice package (v3.16.0) in R (v4.4.2).^59,60^ To impute each culture, we used age, sex, BMI, HIV/CD4 status, smoked substance use (methamphetamine, methaqualone, or cannabis), alcohol use, isoniazid resistance, time to culture positivity at baseline, and all smear positivity and culture results taken before and at the same week as the culture to be imputed, similar to as done previously.^60^ TCC was computed on each of 30 imputed datasets, and model results were pooled across 30 imputations using Rubin’s rules.^59^

### L. Lineage typing

To get lineage calls for the samples, we ran bcftools mpileup and bcftools call to perform variant calling with a ploidy of 1 and a maximum coverage of 200 on the BAM files generated in **Methods Section A.** To further reduce the risk of contaminating reads biasing the lineage typing, we masked 10 genes/regions with high homology to other bacteria: *hsp, clpB, rpoB, rpoC, tuf, rpsC, aspT, rrs, rrl*, and *rrf*. Because mismapping of short reads can lead to false unfixed variants, we additionally masked pre-defined regions of low confidence and sites with an empirical base pair recall < 0.95^61^ and all sites in the *Rv2081c-Rv2082* region (2,338,065-2,340,874).

Finally, we typed lineages using TBtypeR.^62^ This tool identifies *Mtb* lineages and their proportions using a consensus barcode of 10,903 SNVs. The tool can identify 165 distinct lineages, including animal-adapted lineages^63^ and subclades of the L2.2.1 sublineage.^64^ The masking steps above excluded 588/10,903 sites, or 5.4% of the sites used by TBtypeR. Lineage typing using TBtypeR for the culture sequencing samples was performed in the same manner, starting with the BAM files generated in **Methods Section A**.

To identify paired WGS samples for longitudinal analysis, we used all available culture sequencing samples. Among the 452 study participants, 325 had at least two sputum samples sequenced during the first 12 weeks of treatment. We used the TBtypeR lineage calls for further matching. For participants with more than two WGS samples, we first excluded samples with a different lineage from the majority lineage called for that participant’s samples, then we kept the first and last samples for each person to maximize the time interval between samplings. 15 of the 325 participants had single lineages called at both timepoints that disagreed with each other, which could be due to sample mislabeling or undetected mixed infections. These 15 were excluded from longitudinal analyses.

Of the remaining 310 participants, 285 had a single concordant lineage at both time points. We first inspected the mutation rate for fixed (AF > 0.95) mutations in these 285 individuals to determine if it is consistent with previous estimates of the *Mtb* mutation rate. We excluded one participant because their *Mtb* genome gained 17 fixed SNPs, while the others gained 0-3 SNPs each, suggesting that pid T0245 may have mismatched WGS samples. We also excluded pid T0245 from all other longitudinal analyses. We find 19 total variants absent at baseline and fixed at follow-up across 284 individuals. Using a negative binomial model to account for overdispersion caused by different mutation rates across different *Mtb* samples and hosts, we estimate a mutation rate of 0.66 SNPs/genome/year (95% confidence interval = 0.36-1.2), which is consistent with other empirical estimates of the *Mtb* mutation rate.^22–24^

## Supplement

### Results

#### A. Identification of unfixed substitution variants from hybrid assemblies

To build a ground truth dataset of unfixed single nucleotide variants (SNVs), which we define as having a within-sample allele fraction (AF) ≥ 0.05 and AF ≤ 0.95, we performed variant calling using hybrid PacBio / Illumina assemblies as reference genomes. We selected a minimum allele fraction of 5% because variant allele fractions below 5% are more likely to be noise.

A total of 186 PacBio HiFi sequencing samples were available from this cohort. Due to research constraints, not every participant had a bacterial sample sequenced with PacBio reads. These were sequenced from sputum cultures during the first 12 weeks of TB treatment or at 5 months after starting treatment. The reads were assembled according to a previously published pipeline (**Methods**).^65^ Of the 186 long read sequencing samples, 154 were assembled into a single circular contig (**Supplementary Data 2**). These ranged in size from 4.36 to 4.44 megabase pairs (Mbp) with coverages of 14-295x (median = 89.5x) (**Supplementary Fig. 1a**). The lineage (L) distribution is 90 L2, 57 L4, six L3, and one L1.

A total of 767 culture samples from this cohort were sequenced with Illumina sequencing (**Supplementary Data 1**). For unfixed variant detection, we restricted to the 738 out of 767 Illumina samples with an F2 score ≤ 0.03 to focus on changes in monoclonal samples. 172 of the 738 samples were able to be matched to a hybrid assembly from the same participant, though not necessarily from the same culture sample. We verified that the lineage assignments^66^ for the Illumina sample and the hybrid assembly matched.

Illumina reads were aligned to the personal reference genomes, followed by variant calling with freebayes. We chose freebayes because benchmarking multiple variant callers, including those optimized for low-frequency variants, on unfixed variant calling in *Mtb* showed that freebayes achieved the highest precision and recall.^26^ Variants with 0.05 ≤ AF ≤ 0.95 were transferred to H37Rv coordinates using minimap2. We inspected the transfer rate of the unfixed variants for the 172 samples with personal reference genomes, and found that ten variants (9 SNVs, 1 indel) were not transferred between coordinate spaces: five are in the IS6110 transposable element^67^ between H37Rv coordinates 3,710,433 and 3,713,461, and one occurs downstream of this element in the gene *sugI* (coordinates 3,717,090–3,718,598 in H37Rv). However, the start of *sugI* is predicted to be uncertain.^68^ The region around *sugI* is not difficult to map reads to, and we note that this variant was picked up when Illumina reads were aligned to H37Rv. The only step that fails is the transformation of coordinates from the personal genome to H37Rv, which may be complicated by its proximity to the IS6110 element. The other four variants occur in *PPE60* (N = 3) and *PE_PGRS55* (N = 1). Both of these genes demonstrate evidence of recombination with other genes in the *Mtb* genome,^69^ which makes unambiguous coordinate transfer difficult.

To obtain a final list of unfixed variants per sample, we required the coverage at each unfixed variant site to be at least half the global median coverage because short reads from especially repetitive regions can be mismapped, even when using a sample’s own genome as the reference. Finally, all positions within ribosomal RNAs (*rrs, rrl,* and *rrf*), transposable elements, and phage sequences were removed. The ribosomal RNAs are prone to miscalled unfixed variants due to even small amounts of contamination with environmental bacteria because of their high degree of homology across bacteria.^70,71^ A total of 401 unfixed SNVs remained. The number of unfixed SNVs per sample ranged from 0-14, with a median of 3 (**Supplementary Fig. 1b**).

#### B. Unfixed variant calling against the H37Rv reference genome

We used the same alignment and variant calling parameters as for the personal genomes. We filtered for candidate high quality unfixed SNVs using the following criteria: within-sample allele fraction (AF) ≥ 0.05 and AF ≤ 0.95, read depth ≥ 5, mapping quality ≥ 40, at least 2 forward and 2 reverse reads (determined by freebayes) supporting the SNV, and an average Phred quality of bases supporting the variant ≥ 20. We further excluded positions in low mappability regions or with an empirical base pair recall < 0.95,^61^ and all positions in transposable elements, phage sequences, and ribosomal RNAs. The number of unfixed variants per sample begins to increase when F2 (a lineage mixing metric)^72^ > 0.03 (**Supplementary Fig. 1c**), so this is how the threshold was chosen for monoclonal samples. Of the 767 Illumina samples, 738 have an F2 ≤ 0.03. The lineage distribution of the 738 samples is 366 L2, 323 L4, 48 L3, and one L1.

After the above filtering criteria, the number of unfixed SNVs per sample among the 172 samples that also have personal reference genomes ranged from 0-17, with a median of 4 (**Supplementary Fig. 1d**). This is significantly larger than the number of unfixed SNVs per sample determined using variant calling against the personal reference genomes (one-sided paired t-test p = 4.98 x 10^-34^). This indicated that despite the filtering criteria above, many false unfixed SNVs were called when aligning short reads to H37Rv. The range of the difference in the number of unfixed SNVs between variant calling with H37Rv and the personal genomes is 0-14, with an average of 2.7 false unfixed SNVs per sample.

If there is systematic bias in calling unfixed SNVs, we would expect to observe considerable differences in the genes or intergenic regions with the most unfixed SNVs. The genes or intergenic regions with the most common unfixed SNVs are *Rv2082, Rv2319c*, *iniB*, and *uspC*, with 394, 368, 237, and 234 SNVs each, respectively. *Rv2319c* appears to have a structural variant, as evidenced by the high variance in coverage and the large number of discordantly paired reads (green) (**Supplementary Fig. 2a**). We also observed false variants at regions with a large number of reads with soft clipping (**Supplementary Fig. 2b**). In this sample, the variant is fixed, but the reads that do not support the variant are soft clipped, suggesting that they originate from elsewhere in the genome. False unfixed SNVs also occur near fixed indels. Reads that do not sufficiently cover an indel do not unambiguously provide evidence for the indel, and instead, they appear to support one or more substitutions (**Supplementary Fig. 2c**).

#### C. Duplication of *Rv2081c-Rv2082* causes numerous false unfixed SNVs

Among samples with both long and short read sequencing, when the Illumina reads were aligned to H37Rv, we observe 56 unfixed SNVs in the gene *Rv2082* between coordinates 2,338,773 and 2,340,477 (**Supplementary Fig. 2d**) across 8 samples (6 L3 and 2 L4 samples, but the two L4 samples come from a single individual). None of these was found when aligning Illumina reads to the personal genomes for these samples. Suspecting a structural variant here, we aligned PacBio reads to H37Rv and performed structural variant detections with sniffles. Sniffles identifies 2.6 or 5.3 kilobase pair (kbp) insertions in this region, as evidenced by the large increase in coverage between positions 2,338,129 and 2,340,759 in H37Rv, which covers most of *Rv2081c* and *Rv2082* (**Supplementary Fig. 3c**).

We extracted the regions between the start of *Rv2081c* and the beginning of *Rv2083* for the 8 samples. The lengths of these regions are 5,466 base pairs for the L3 samples and 8,111 base pairs for the L4 samples. We then performed individual pairwise alignments between the 8 sample sequences to H37Rv. Based on the aforementioned lengths, we suspected that there are two extra copies of this region in L4 and one extra copy in L3. Therefore, we concatenated the H37Rv sequence to itself twice before aligning to the L3 samples and concatenated the H37Rv sequence to itself thrice to align it to the L4 samples. For the L3 samples, the alignments are all 5,633 nucleotides with 5,097 (90.5%) exact matches. For the L4 samples, the alignments are of length 8,380 nucleotides with 7,626 (91.0%) exact matches. This suggests that there is one additional paralog in L3 and two additional paralogs in L4. The L4 samples belong to L4.1.2, and they are the only samples of this L4 sublineage with PacBio sequencing that have this duplication. The large number of SNVs within the duplicated regions indicate sequence divergence between the paralogs of this region or mismapping due to the repetitive nature of this region (**Supplementary Fig. 3a-b**).

To investigate this duplication in the rest of the short-read sequencing sample set, we called structural variants from short read sequencing alignments to H37Rv using delly. We identified structural variants with length ≥ 2,500 base pairs and at least 10 paired end reads of support in 34 participants’ baseline samples, all of which are in lineages 3 and 4. The sublineage distribution of lineage 4 samples is four L4.1.2, one L4.2.2, and one L4.1 (**Supplementary Fig. 3d**). L4.1.2 and L4.2.2 are monophyletic, while the L4.1 sample is farther away. The clustering of this duplication in phylogenetic space suggests that it arose multiple times in different subclades and then was propagated through transmission. A larger long read sequencing sample set with a wider lineage distribution (in particular, L1 and L7) is needed to develop hypotheses for the phylogenetic history of this duplication.

#### D. Logistic regression model to predict the accuracy of an unfixed SNV

Given the observations about reference bias-induced false positive unfixed variant calls, we built a logistic regression model to predict the probability of a candidate unfixed SNV being accurate using the following features:

● **Discordant reads:** Number of discordantly paired reads normalized to coverage
● **Base quality:** Average base quality of bases supporting the variant. Most reads at the site of an unfixed variant support the reference allele, so the average base quality computed by variant callers reflects the qualities of both reference and alternate alleles.
● **Coverage:** Ratio of coverage at the site to the rolling average
● **Strand bias:** Absolute value of the difference between 0.5 and the proportion of reads supporting the variant that are in the forward orientation
● **Soft clipped bases:** Number of soft clipped bases at the variant site normalized to coverage
● **Soft clipped reads:** Ratio of reads supporting a variant that are soft clipped to the total number of reads supporting the variant. This is different from the above variable because reads can be soft clipped elsewhere, not just at the site with the variant.
● **Intra-read variant position:** Median of the normalized position within reads that support the variant. The closer the variant is to the edge of the read, the smaller this value is.

The coverage rolling average was computed with a window size of 100 base pairs, and we took the maximum of the rolling averages computed from the up-and downstream directions. Because we expected that increases and decreases in coverage have different effect sizes on the probability of a variant being real (*i.e.,* a drop in coverage is likely more indicative of a false unfixed SNV than an increase in coverage), we split the coverage ratio variable into two variables, one for ratios ≥ 1 and one for ratios < 1.

After first excluding all variants in *Rv2081c* and *Rv2082* and the intergenic region, which spans positions in the range [2338065, 2340874] in H37Rv, the model was trained on 802 candidate unfixed SNVs (401 real, 401 not real) from the 172 WGS samples with matched personal reference genomes. The variants and associated statistics were derived from variant calling against H37Rv, and the real labels were derived from their presence or absence when variant calling with the personal reference genomes.

Because the model is focused on accuracy for this dataset, rather than generalizability, we trained a single model on all 802 variants. Predictions were then obtained for 2,366 additional unfixed SNVs from 565 WGS samples without personal reference genomes. Five of the eight estimated odds ratios were significant at a significance level of *α*=0.05, and the effect sizes were consistent with expectation, with odds ratios > 1 for base quality and intra-read variant position, and odds ratios < 1 for the others (**Supplementary Table 3**). Of the 2,366 candidate unfixed SNVs in the 565 WGS samples without personal reference genomes, 1,205 were predicted to be real (**Supplementary Data 4**). On the train set, the classification statistics are as follows: area under the curve (AUC) = 0.991, precision = 0.978, and recall = 0.978. There are 9 false negatives and 9 false positives.

The distributions of the number of unfixed SNVs per sample detected from the personal reference genomes and detected from H37Rv are not significantly different by a two-sided Kolmogorov-Smirnov test (p = 0.96), while the distributions were significantly different (p = 3.9 x 10^-23^) before filtering the H37Rv-called unfixed SNVs using the logistic regression model. 224/738 (30%) samples of the full dataset and 45/172 (26%) samples of the ground truth dataset have no unfixed SNVs, while only 38/738 (5.1%) of samples had 0 unfixed SNVs before filtering.

#### E. Adjusting indel allele fractions to exclude false positives

Calling unfixed indels is generally less affected by reference bias because variant callers require greater evidence to call indels than SNVs. The most salient issue in accurately calling low frequency indels is reads not sufficiently covering an indel, especially in repetitive regions, which are common in the *Mtb* genome. These reads artificially bring down the allele fraction of the indel, making it appear as though a fixed indel is unfixed. We excluded reads found too close to the candidate indel and reads with soft clipping because these are unable to support an indel, even if it is fixed (**Methods**). After running the pipeline, each indel has an adjusted allele fraction computed using only reads that sufficiently span the indel site.

We performed similar benchmarking using the 172 available hybrid assemblies. We compared unfixed indels, those with 0.05 ≤ AF ≤ 0.95 called using H37Rv as the reference genome against those called using the personal assemblies as individual references. A single unfixed indel was not able to be transferred from a personal genome to H37Rv coordinates, but it occurs 4 base pairs upstream of the IS6110 element between *Rv3324c* and *Rv3328c*, where pairwise genome alignment is ambiguous. A total of 67 true indels were called across the 172 samples with matched personal genomes. The number of indels per sample ranges from 0-4 with an average of 0.39. A total of 2,724 unfixed indels were called from these samples after variant calling against H37Rv (67 true positives and 2,657 false positives). After adjusting the AFs and excluding indels with an AF > 0.95, only 3 false positives remained.

After aligning short reads to H37Rv and performing the same quality control as in **Methods Section B,** 6,342 unfixed indels were identified in the full set of 738 WGS samples. The number of unfixed indels ranged from 0-18 with an average of 8.6 per sample, which is significantly larger than the distribution determined from the personal reference genomes (p = 0, Welch’s t-test). After running the above pipeline on the full dataset of 738 samples, there are only 278 total unfixed indels (**Supplementary Data 7**). The number of unfixed indels ranges from 0-5, with an average of 0.38 unfixed indels per sample, which is not significantly different from the distribution of unfixed indels determined on the personal reference genomes (p = 0.42, two-sided Welch’s t-test). There is no significant difference in the number of unfixed indels per sample across the four lineages (p = 0.32, likelihood ratio test). Two-thirds of the unfixed indels are frameshifts (N = 186). The remainder consists of 40 inframe deletions, 17 inframe insertions, and 35 intergenic variants.

#### F. Additional unfixed variant burden associations

In Kaplan Meier analysis, we observed a non-monotonic relationship between baseline pathogen diversity and the risk of outcomes (**Fig. 4b**). A boundary knot of 3 was found to minimize the Akaike Information Criterion (AIC) (**Supplementary Fig. 5a**). We therefore fit different hazard ratios below and above an unfixed variant count of 3 in Cox proportional hazards models. Adding unfixed variant burden at baseline to the multivariate model increases the concordance index from 0.68 to 0.73, reduces the AIC from 278.1 to 276.0, and significantly increases the log-likelihood (likelihood ratio test p-value = 0.01). The Cox-Snell pseudo R^2^, indicates that unfixed variant burden explains an additional 1.7% in the variation in outcome over the base model with patient covariates only.

#### G. Unfixed variant burden at baseline is a better predictor of composite outcomes than unfixed variant burden at follow-up

To compare model fits between unfixed variant burdens at baseline and follow-up, we performed the analysis on the 262/364 individuals with longitudinal sequencing. We used boundary knots of 1 for baseline and 2 for follow-up to maximize the AIC in each case (**Supplementary Fig. 5c-d**). Unfixed variant burden at baseline increases the concordance index from 0.72 to 0.77, lowers the AIC from 216.5 to 212.5, and explains an additional 3.0% of the variation in outcome time, whereas unfixed variant burden at follow-up has concordance index = 0.74 and AIC = 216.2 and explains 1.6% of additional variation. These results demonstrate that the relationship between unfixed variant burden and composite outcomes is robust to the sampling time when samples are taken within the first 12 weeks since treatment initiation.

#### H. Unfixed variant burden does not associate with time to culture conversion

We additionally tested if unfixed variant burden associates with time to negative culture conversion (TCC). Sputum was sampled weekly for the first 12 weeks of treatment. TCC was defined as the number of weeks between the time of treatment initiation and the first of two consecutive *Mtb*-negative sputum cultures not followed by another positive culture. To be included in the TCC analysis, participants had to have at least 3 uncontaminated sputum samples and a positive *Mtb* culture within the first 5 weeks of initial screening, leaving 323/364 patients remaining for the TCC analysis. The median TCC is 9 weeks.

The TCC Cox proportional hazards model was stratified by 4 patient covariates – previous TB disease, underweight (*i.e.,* BMI < 18), smear positivity, and PLI > 25% (**Supplementary Fig. 6a-d**) – in order to satisfy the Cox proportional hazards assumption. The directions of these effects generally agree with expectation, and the log-rank test indicates significantly different Kaplan-Meier (KM) estimates, with previous TB (p = 0.11), underweight (p = 0.017), smear positivity (p = 3.3 x 10^-8^), and PLI > 25% (p = 6.6 x 10^-5^) being associated with longer TCC. After these stratifications, only age and TTP at baseline are associated with longer TCC. Unfixed variant burden is not associated with TCC (**Supplementary Fig. 6e**).

#### I. Mixed infections are not associated with worse outcomes or TCC

However, in this dataset, mixed infections, either represented by F2 score or binarized at 0.03, were not associated with worse outcomes over clonal infections. In univariate models, F2 score had an aHR of 1.19 (95% CI 0.64 - 2.20, p = 0.58), and the binary variable F2 > 0.03 had an aHR of 1.54 (95% CI 0.36 - 6.50, p = 0.56). The prevalence of mixed infections at baseline is low (5%), and therefore the dataset may be underpowered to detect an association if it exists.

## Data

**Supplementary Data 1: 767 Illumina sequencing samples and sequencing information**. Each sample (denoted by its MFS-ID in the “SampleID” column) has the associated participant ID (“pid”), a more descriptive sample ID (“Original_ID”), timepoint (“Sampling_Week”), and statistics about contamination, sequencing depth, lineage mixing, and lineage classification. The lineage classifications were determined by TBtypeR.^62^ “mix_phylotypes” is a comma-separated list of the sublineages; “mix_props” is a comma-separated list of the proportions that each lineage in “mix_phylotypes” composes, in the same order; and “n_phy” is the total number of lineages detected at ≥ 1%. Each value in “Original_ID” is composed of the participant ID (with an “S” instead of a “T”) and the sampling week. Timepoint suffixes of “m5” and “PM12” indicate month 5 since enrollment and post-treatment month 12, respectively.

**Supplementary Data 2: Hybrid assembly characteristics and short-read samples matched to hybrid assemblies.** Sheet 1: 154 complete, high-quality *Mtb* assemblies generated from PacBio HiFi sequencing and polished with matched Illumina sequencing. Lineage classifications made by fast-lineage-caller.^73^ Sheet 2: 179 short-read samples matched to the 154 assemblies in Sheet 1. All the short-read samples are matched to assemblies from the same participant, but not necessarily from the same timepoint. “SampleID” and “Original_ID” refer to the short-read sample. “ASM_SampleID” and “ASM_Original_ID” are the short-read and long-read samples used to generate the hybrid assembly in “ASM.” One short-read sample from the same participant and same timepoint was sequenced twice: MFS-635 and MFS-878.

**Supplementary Data 3: Baseline and longitudinal samples used for analyses.** Sheet 1: 395 participants with baseline short-read WGS samples (within the first 2 weeks of enrollment) and F2 ≤ 0.03. Sheet 2: 310 participants, each with two short-read WGS samples. 285 of the participants have identical lineages and F2 ≤ 0.03 for both samples; these participants have a value of 1 in the “PassQC” column. 25 participants have mixed lineage samples for at least one timepoint; these participants have a value of 0 in the “PassQC” column.

**Supplementary Data 4: Training and new data for the logistic regression error model.** Sheet 1: 802 variant calls for the training data with freebayes VCF metadata, the features passed into the error model, and a column “Real” for whether or not the variant call was real and also detected from the personal genome. Sheet 2: 2,366 variant calls from the samples without personal reference genomes. Each variant call has freebayes VCF metadata, the features for the error model, and columns for the predicted probability that the variant call is real (“predicted” column) and a predicted binary classification of real or not real using the selected threshold of 0.67 (“pred_class” column).

**Supplementary Data 5: Unfixed and Fixed Variant Calls for all samples.** Sheet 1: Unfixed variant calls for 285 longitudinal WGS samples. Sheet 2: Unfixed and fixed variant calls for 285 longitudinal WGS samples. Sheet 3: Unfixed variant calls for 395 baseline WGS samples. Sheet 4: Unfixed and fixed variant calls for 395 baseline WGS samples. Sheet 5: Unfixed variant calls for all 738 WGS samples. Sheet 6: Unfixed and fixed variant calls for all 738 WGS samples.

**Supplementary Data 6: Enrichment results of unfixed substitutions.** Unfixed variant enrichment results across regions (genes or intergenic regions) for all baseline (Sheet 1) and follow-up (Sheet 2) WGS samples. For both sheets, p-values for synonymous and non-synonymous mutations are given separately, along with p-values for the combined set. Both original and Bonferroni-corrected p-values are given.

**Supplementary Data 7: Unfixed indels.** All 278 unfixed indels identified across 738 WGS samples, including phase variant annotations. A phase variant is defined as an insertion or deletion of one unique nucleotide from a repeated sequence of the same nucleotide at least 7 times.

---

**Supplementary Figure 1.**
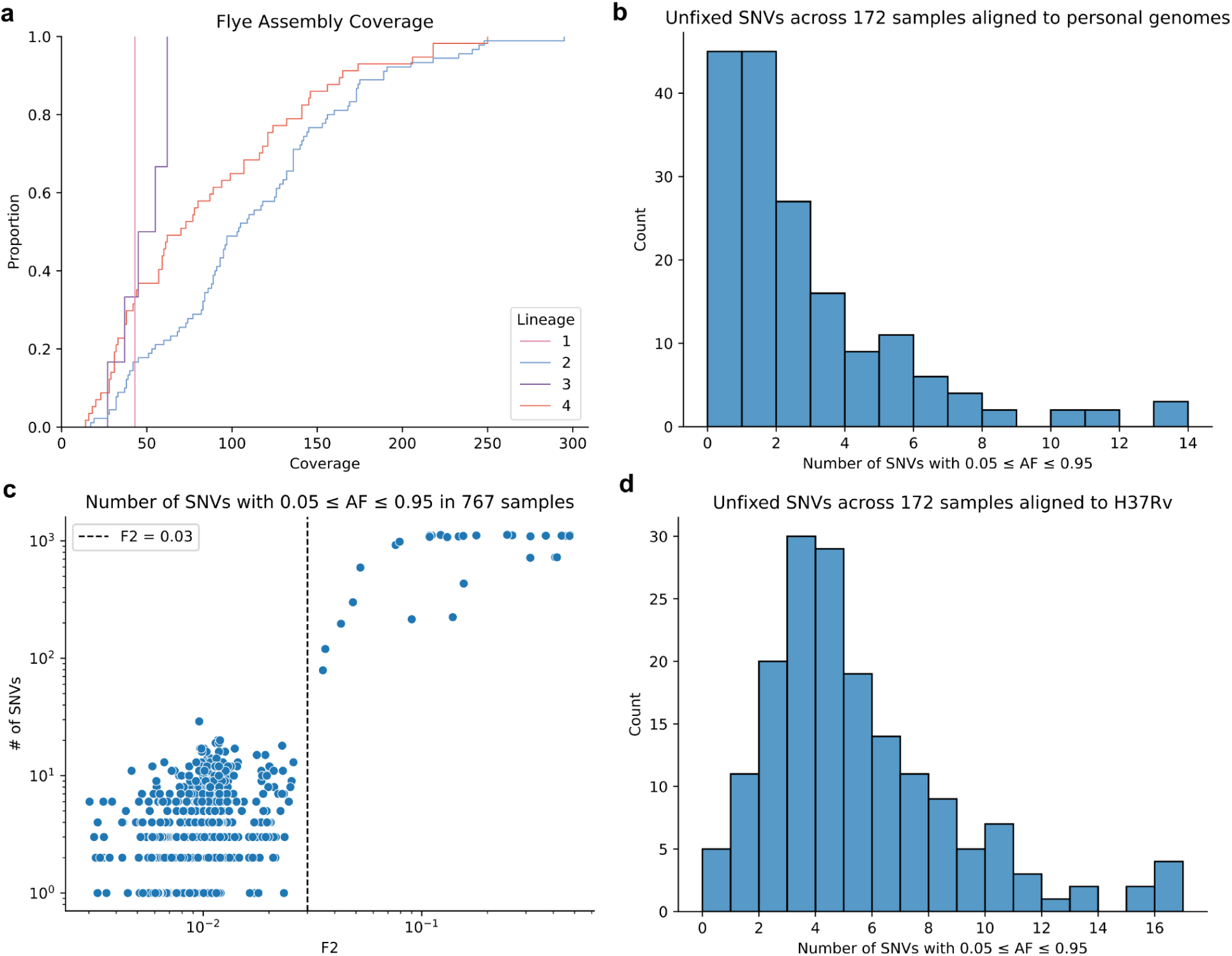
Hybrid assembly coverage statistics and comparison of unfixed SNVs called from personal reference genomes and H37Rv. **a**: Assembly coverage based on Flye of 154 high quality hybrid assemblies. **b**: Distribution of number of unfixed single nucleotide variants (SNVs) per sample for 172 Illumina samples aligned to 154 personal reference genomes. **c**: Number of unfixed SNVs per sample vs. F2 strain mixing metric for all 767 Illumina samples aligned to H37Rv. **d**: Distribution of number of unfixed SNVs per sample for 172 samples in panel **b** but variants called against H37Rv. The total number of SNVs in panel **b** is 401, and the total number of SNVs in panel **d** is 866.

**Supplementary Figure 2.**
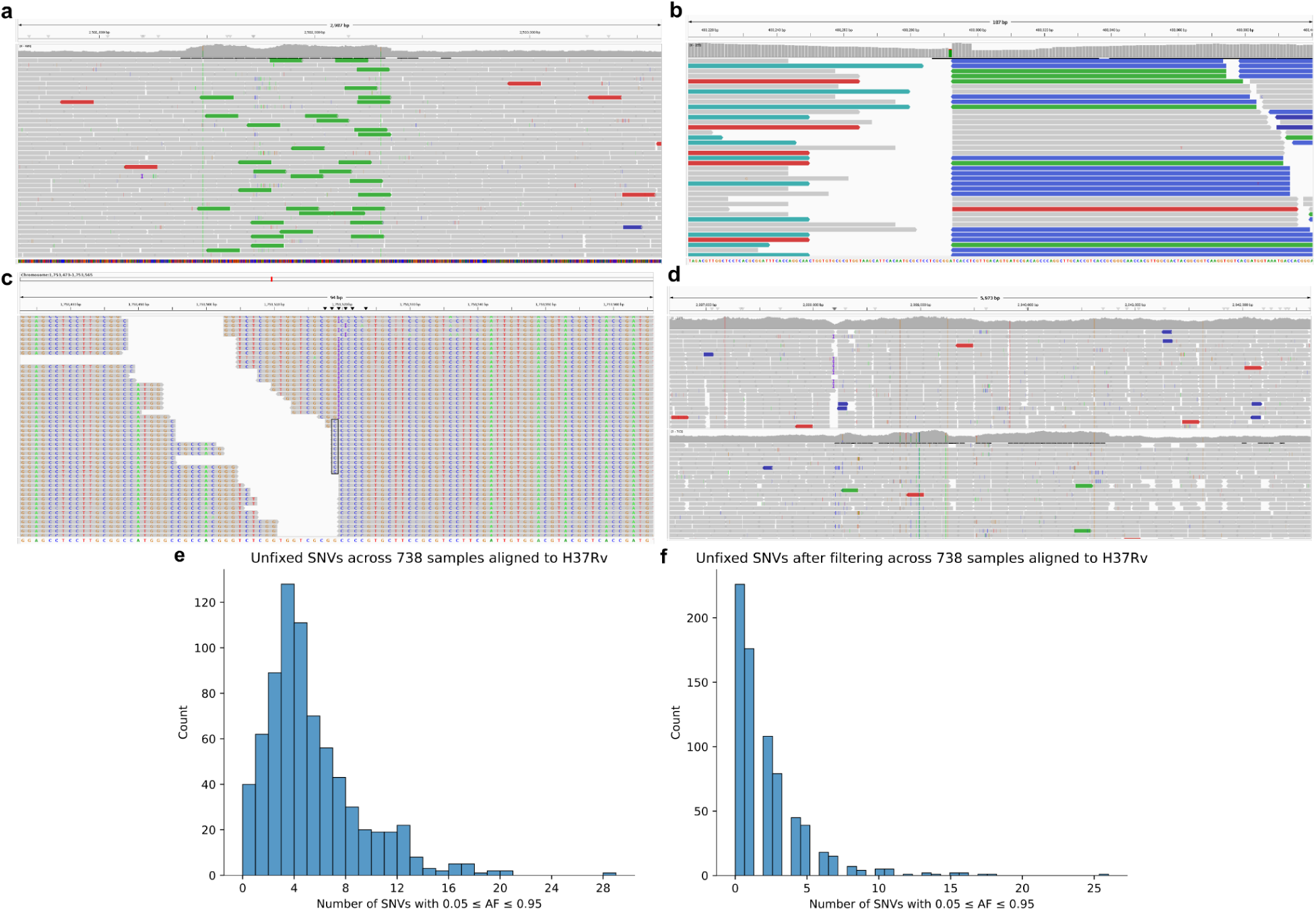
Examples of false positive unfixed variants used to determine relevant alignment statistics. **a-c**: Examples of alignments of Illumina reads to H37Rv, showing reference bias-associated problems of discordantly paired reads (**a**), soft clipping (**b**), and proximity to indels (**c**). In **c**, the reads that support a G>C substitution just before the indel are boxed. **d**: Alignments of Illumina reads to H37Rv for the gene *Rv2082* for two samples: the top sample is L2.2.1, and the bottom sample is L3. The L3 sample shows a large number of unfixed SNVs in a region of increased coverage. **e**: Distribution of number of unfixed SNVs per sample for 738 samples before logistic model filtering. **f**: Distribution of number of unfixed SNVs per sample for 738 samples after all filtering and exclusion steps.

**Supplementary Figure 3.**
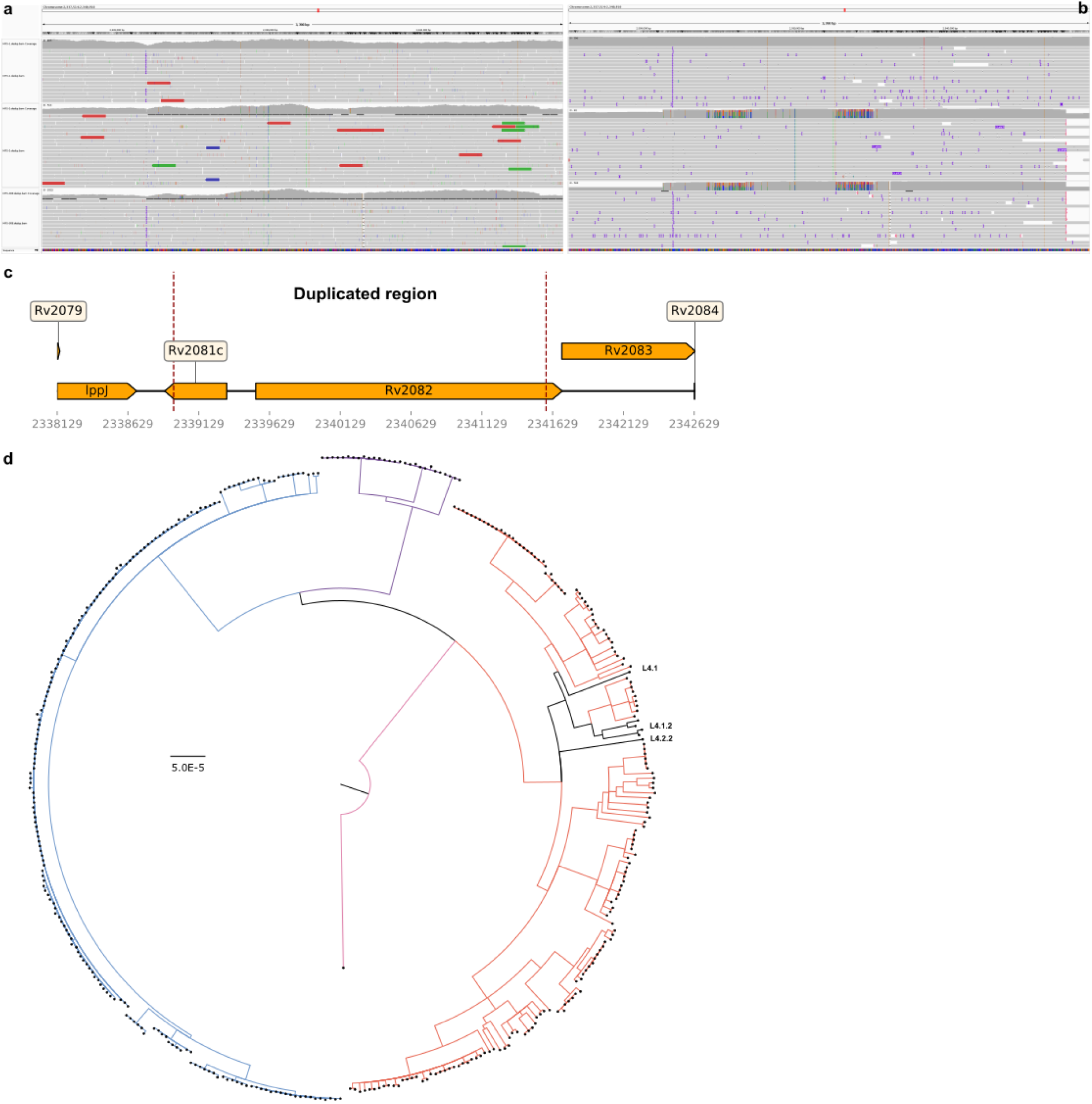
*Rv2081c-Rv2082* duplication in lineage 4 sublineages and lineage 3. **a**: Short reads aligned to the H37Rv reference genome for L2 (top), L3 (middle), and L4 (bottom) samples. **b**: Long reads aligned to H37Rv for the same three samples in the same order as in **a. c:** Schematic of the duplicated region in H37Rv. The dashed red lines are the boundaries of the duplication. **d**: Maximum likelihood tree of 395 Illumina samples constructed using 14,049 genome-wide single nucleotide variants. Red = L4, purple = L3, blue = L2, pink = L1. All 28 L3 samples and 6 L4 samples have the duplication. The 6 L4 samples with the duplication are colored in black and annotated on the right side of the tree.

**Supplementary Figure 4.**
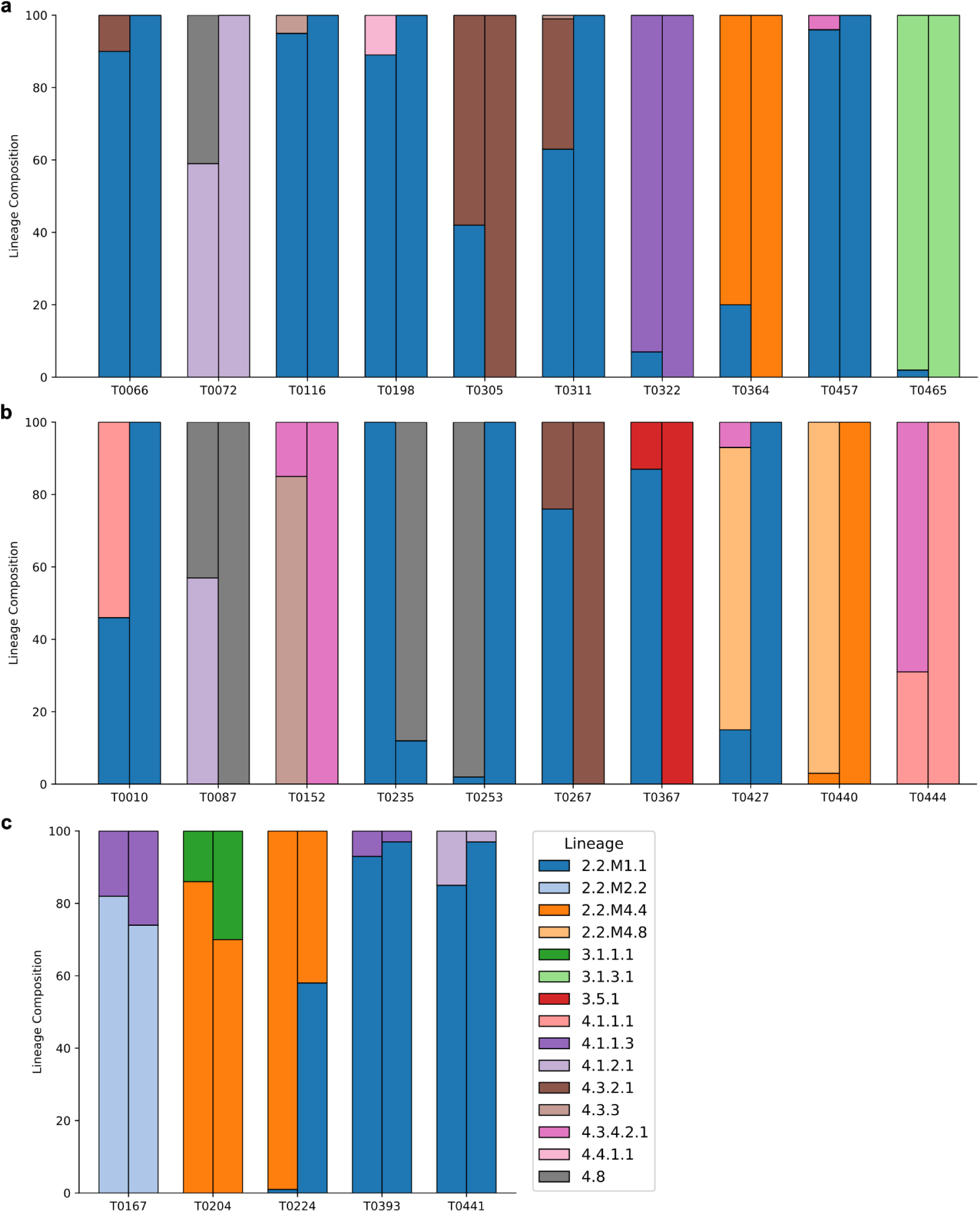
Lineage fraction changes for 25 participants with mixed infections at baseline or follow-up. a: 10 participants in whom the major lineage at baseline remained the major lineage at follow-up. **b:** 9 participants in whom the minor lineage at baselinebecame the major lineage at follow-up and one participant (T0235), who had a single lineage at baseline, which became the minor lineage at follow-up. **c:** 5 participants who had mixed infections at both baseline and follow-up. The lineage legend is the same for all three panels.

**Supplementary Figure 5.**
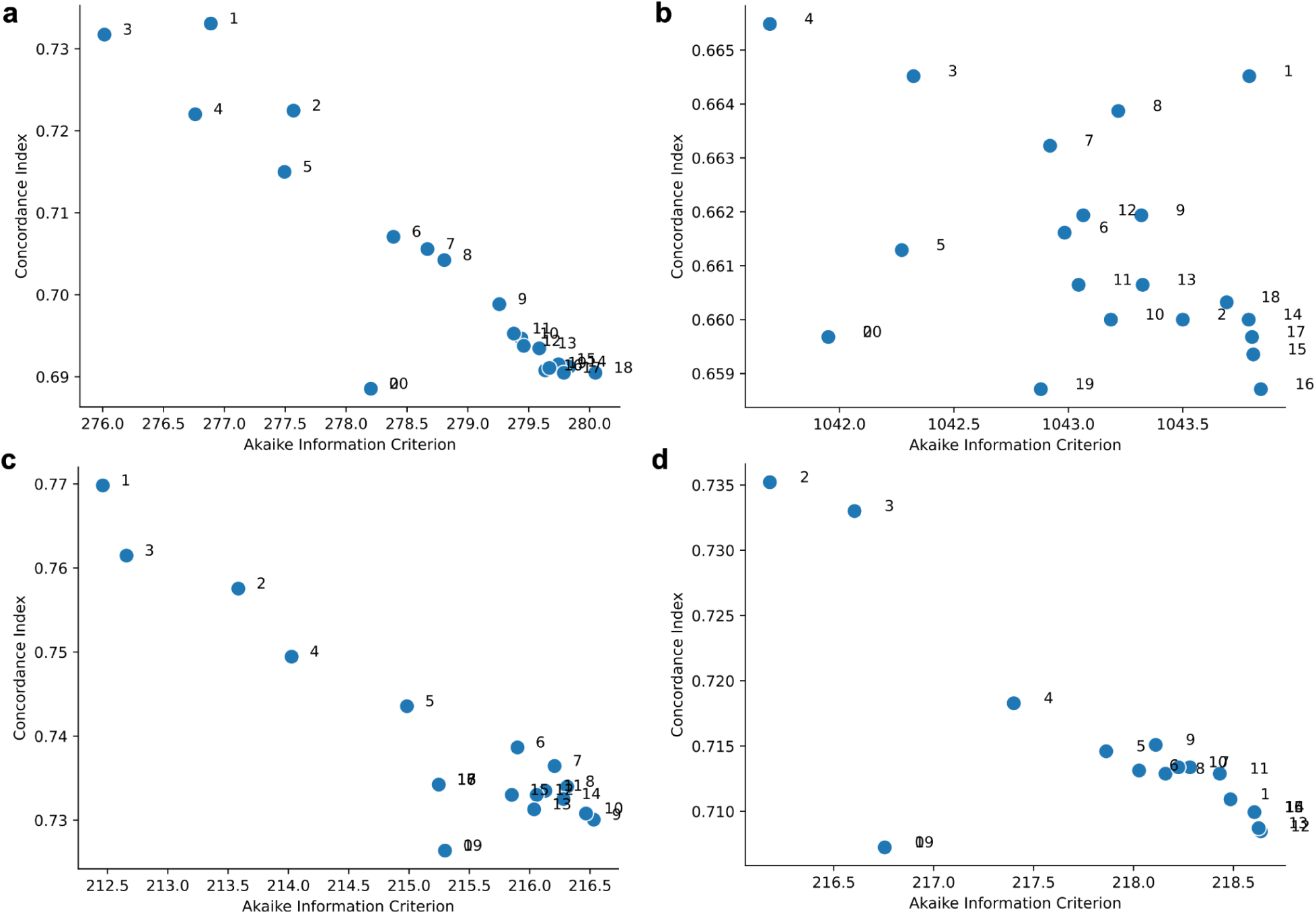
Scatterplots of Concordance Index vs. Akaike Information Criterion to select boundary knots for unfixed variant burden splines. Different panels are different multivariate Cox proportional hazards models with unfixed variant burdens at either baseline or follow-up. **a:** Unfixed variant burden at baseline in a Cox model predicting time to unfavorable outcome for 364 participants. **b:** Unfixed variant burden at baseline in a model predicting time to culture conversion for 323 participants. **c:** Unfixed variant burden at baseline in a Cox model predicting time to unfavorable outcome for 262 participants with longitudinal sequencing. **d:** Unfixed variant burden at follow-up in a Cox model predicting time to unfavorable outcome for 262 participants with longitudinal sequencing.

**Supplementary Figure 6.**
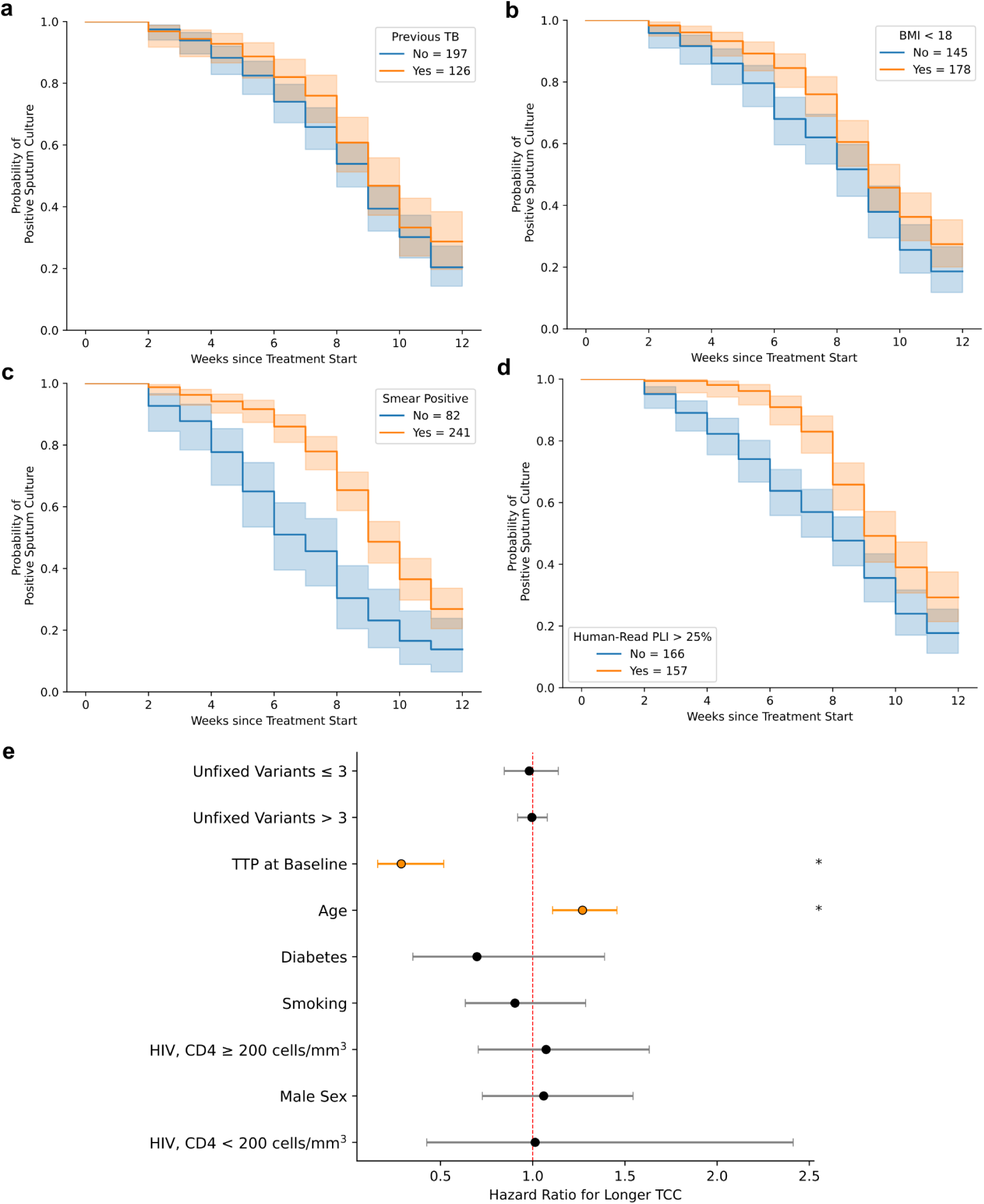
Cox proportional hazards model for time to culture conversion (TCC) for 323 participants with clonal. *Mtb* **infections**. **a-d**: Kaplan-Meier estimates stratified by 5 variables to satisfy the proportional hazards assumption. In order, previous TB **(a)**, underweight (BMI < 18) **(b)**, smear positivity **(c)**, PLI > 25% **(d)**. **e**: Forest plot of hazard ratiosbetween TCC, patient covariates, and unfixed variant burden at baseline, with 95% confidence intervals (Wald). Standard errors were computed by the Cox regression fitter in lifelines. Variables with two-sided p-values < 0.05 are shown in orange and denoted by an asterisk. All reported hazard ratios are associations with longer TCC. The hazard ratio for age is per 10 year increase, the hazard ratio for unfixed variant burden is per 10 variant increase.

**Supplementary Table 1.**
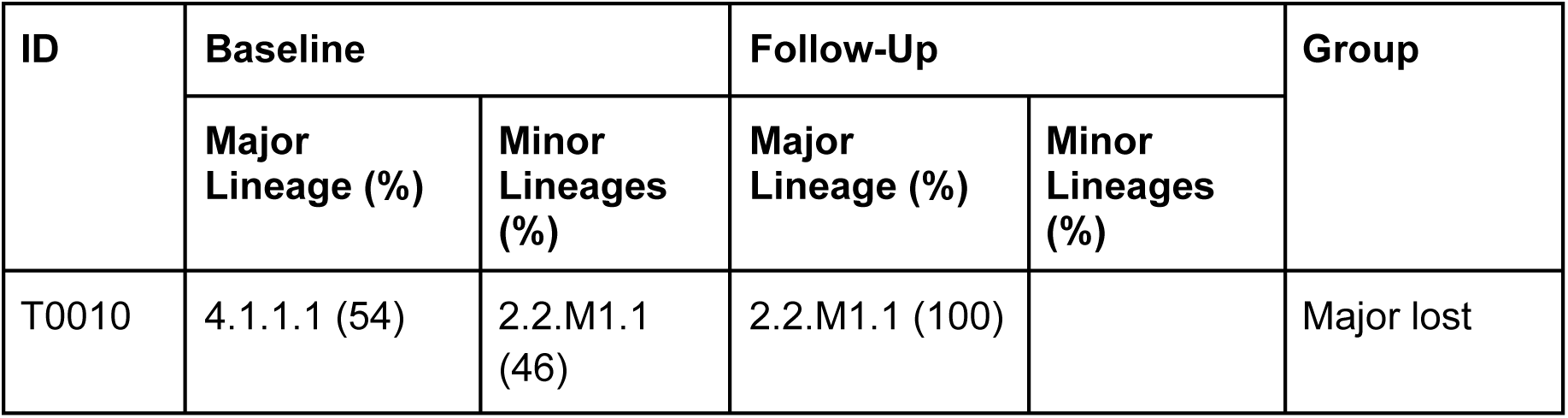
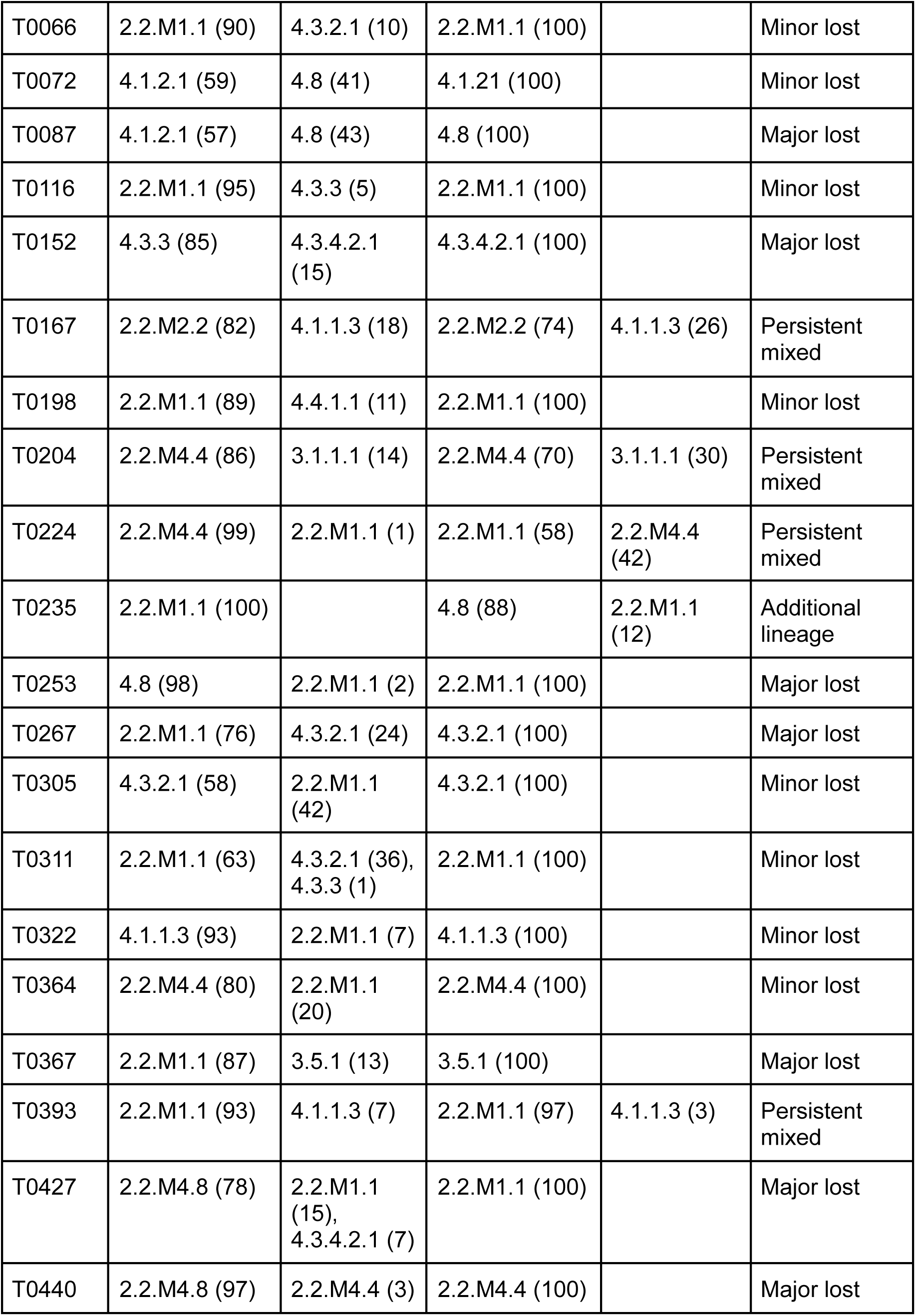
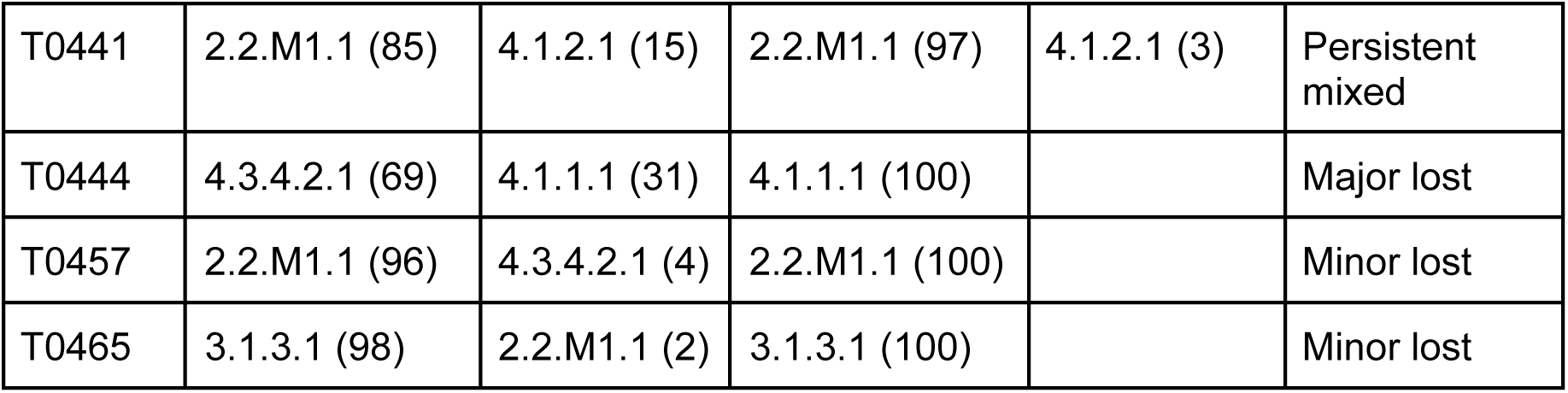
Lineage proportions of culture samples of 25 participants with mixed infections at either baseline or follow-up. Mixed infections are those with multiple lineages detected with a prevalence of at least 1% by the TBtypeR tool.^62^ Both timepoints occur during the first 12 weeks of enrollment. Only one of these participants had a sequenced culture at month 5: pid T0204 had a 100% L2.2.M4.4 sample then. The participants are grouped into categories describing changes relative to baseline. All lineage 2 samples in this cohort are part of the L2.2.1 sublineage, which is further divided into subclades.^64^

**Supplementary Table 2.** Within-sample allele frequencies of known resistance mutations in participants with mixed infections. The two variants in *rpsL* are associated with streptomycin resistance, and the other four variants are associated with isoniazid resistance.^2^ All four individuals had phenotypically measured isoniazid resistance at baseline. Phenotypic streptomycin resistance was not measured in this cohort.

| ID | Variant | Baseline |  | Follow-Up |  | Interval (weeks) |
| --- | --- | --- | --- | --- | --- | --- |
|  |  | AF | Lineages | AF | Lineages |  |
| T0116 | inhA_c.-154G>A | 92% | 2.2.M1.1 (95%),<br>4.3.3 (5%) | 100% | 2.2.M1.1 | 5 |
| T0305 | inhA_p.Ser94Ala | 56% | 4.3.2.1 (58%),<br>2.2.M1.1 (42%) | 100% | 4.3.2.1 | 7 |
| T0322 | katG_p.Ser315Thr | 96% | 4.1.1.3 (93%),<br>2.2.M1.1 (7%) | 100% | 4.1.1.3 | 4 |
|  | rpsL_p.Lys43Arg | 93% |  | 100% |  |  |
| T0441 | inhA_c.-777C>T | 84% | 2.2.M1.1 (85%),<br>4.1.2.1 (15%) | 96% | 2.2.M1.1 (97%),<br>4.1.2.1 (3%) | 7 |
|  | rpsL_p.Lys88Arg | 86% |  | 97% |  |  |

**Supplementary Table 3.** Odds ratios and p-values in the logistic regression model. . Variables are ordered by increasing p-value. Confidence intervals are Wald intervals.

| Variable | p-value | Odds Ratio (OR) | 95% confidence interval of OR |
| --- | --- | --- | --- |
| Intra-read position (closer to middle) | $4.24 \times 10^{-14}$ | 11.1 | 5.93 – 20.6 |
| Discordant reads | $3.58 \times 10^{-10}$ | 0.003 | $4.91 \times 10^{-4} - 0.018$ |
| Soft clipped bases | $2.31 \times 10^{-9}$ | $6.37 \times 10^{-48}$ | $2.09 \times 10^{-63} - 1.94 \times 10^{-32}$ |
| Strand bias | $2.29 \times 10^{-5}$ | 0.325 | 0.193 – 0.546 |
| Soft clipped reads | 0.013 | 0.415 | 0.208 – 0.828 |
| Average base quality | 0.094 | 1.60 | 0.924 – 2.75 |
| Coverage increase | 0.110 | 0.744 | 0.518 – 1.07 |
| Coverage decrease | 0.274 | 0.452 | 0.109 – 1.87 |

**Supplementary Table 4.** Empirically estimated unfixed mutation rates for each substitution at baseline (BL) and follow-up (FU) . The mutations are ordered by decreasing rate in baseline samples. The ratio of average transition rate to average transversion rate is3.54 at baseline and 4.20 at follow-up. These values were used to determine the expected rate of each type of substitution to parameterize the Poisson test.

| Mutation | Type | Rate in BL Samples | Rate in FU Samples |
| --- | --- | --- | --- |
| C > T | Transition | $1.17 \times 10^{-4}$ | $1.04 \times 10^{-4}$ |
| A > G | Transition | $1.15 \times 10^{-4}$ | $9.49 \times 10^{-5}$ |
| G > A | Transition | $1.11 \times 10^{-4}$ | $9.07 \times 10^{-5}$ |
| T > C | Transition | $1.07 \times 10^{-4}$ | $1.07 \times 10^{-4}$ |
| A > C | Transversion | $5.93 \times 10^{-5}$ | $4.09 \times 10^{-5}$ |
| C > A | Transversion | $4.41 \times 10^{-5}$ | $2.97 \times 10^{-5}$ |
| T > G | Transversion | $4.22 \times 10^{-5}$ | $3.30 \times 10^{-5}$ |
| G > T | Transversion | $3.53 \times 10^{-5}$ | $3.05 \times 10^{-5}$ |
| C > G | Transversion | $3.10 \times 10^{-5}$ | $2.28 \times 10^{-5}$ |
| G > C | Transversion | $2.28 \times 10^{-5}$ | $1.94 \times 10^{-5}$ |
| A > T | Transversion | $1.19 \times 10^{-5}$ | $6.59 \times 10^{-6}$ |
| T > A | Transversion | $6.59 \times 10^{-6}$ | $3.96 \times 10^{-6}$ |

**Supplementary Table 5.** Changes in unfixed indels in homopolymeric tracts of a single nucleotide repeated at least 7 times. The change in allele frequency of each variant is given in the “AF Change” column. Multiple values are in different individuals. The allele fractions of variants present below the lower limit of 5% were manually estimated from the alignment. The time between paired WGS samples for each participant in the table ranges from 5 to 7 weeks. With the exception of the variants in *Rv0759c, espR,* and *Rv1190*, all other variants occur in a single participant each. Because there are 12 samples (11 participants) with variants in *Rv0759c*, individual allele fractions are not listed. The individual with the *frdB* variant only had a baseline sample sequenced.

| Region | Function | Indel | AF Change | Lineages |
| --- | --- | --- | --- | --- |
| Upstream of *Rv0759c* | Unknown | 854,252 delC<br>854,252 insC | Many | 2, 4 |
| *Rv1190* | Unknown | 1,333,661 insG<br>1,333,661 insG<br>1,333,661 insG<br>1,333,661 insGG<br>1,333,661 insGG<br>1,333,661 insGG | 0 → 28%<br>0 → 8%<br>0 → 15%<br>95 → 0%<br>88 → 0%<br>90 → 97% | 2 |
| Upstream of *epsR* | ESX-1 transcriptional regulator of *espACD* | 4,323,354 insG | 71 → 99%<br>32 → 99% | 2 |
| *PE_PGRS14* | Unknown | 928,285 delC | 0 → 7.7% | 3 |
| *glpK* | Glycerol kinase | 4,139,183 insC | 6.3 → 0% | 4 |
| *Rv0954* | Cell division | 1,066,020 delG | 1.3 → 6.4% | 4 |
| *espK* | ESX-1 protein | 4,358,979 insG | 5.6 → 0% | 3 |
| Upstream of *Rv1990A* | Dehydrogenase, possible pseudogene | 2,234,247 insG | 7.3 → 0% | 2 |
| *aspB* | Aminotransferase | 4,007,271 delG | 73 → 0% | 4 |
| *frdB* | Reductase | 1,760,164 insGGG | 86% at baseline | 2 |
| *Rv1894c* | Unknown | 2,141,408 insG | 0 → 9% | 2 |

